# Childhood trauma, accelerated biological aging and chronic pain across two population cohorts

**DOI:** 10.64898/2026.09.08.26362416

**Authors:** Hannah Russell, Sam Singleton, Lesley A. Colvin, Gary J. Macfarlane, Steve Cole, Christopher G. Bell, Tim G. Hales

**Author notes:** **Corresponding author:** Tim G. Hales. Division of Neuroscience, School of Medicine, Ninewells Hospital, University of Dundee, Dundee, UK.

## Abstract

**Background:** Adverse childhood experiences are associated with reduced healthy longevity, multimorbidity, and chronic pain, but the biological mechanisms involved remain poorly understood. Epigenetic age acceleration (EAA), a DNA methylation marker of biological aging, may represent a pathway through which early-life stress becomes embedded. We therefore examined associations between childhood trauma (CT), EAA and chronic pain, and investigated whether accelerated biological aging mediates the relationship between CT and chronic pain in women and men.

**Methods:** We analysed two independent population cohorts from the UK and USA: Generation Scotland (n = 901) and MIDUS (n = 1041). EAA was derived from first-generation (Hannum and Horvath) and second-generation (GrimAge and PhenoAge) DNA methylation clocks relative to chronological age. DunedinPACE, a third-generation measure, was used to estimate pace of aging. Associations between CT, EAA and chronic pain were examined using sex-stratified regression models adjusted for confounders and covariates, followed by mediation analyses.

**Findings:** CT was associated with chronic pain and with accelerated biological aging measured primarily using second- and third-generation clocks. Associations were consistently stronger in women, and smoking-related methylation contributed substantially to the relationship between EAA and chronic pain. Mediation analyses revealed that EAA partially accounts for the link between CT and chronic pain in women.

**Interpretation:** Second- and third-generation clocks appeared more sensitive to the long-term consequences of childhood trauma than clocks trained primarily on chronological age. The findings suggest that CT contributes to chronic pain vulnerability partly through accelerated biological aging pathways, particularly in women.

**Funding:** UK Research and Innovation, Lily, and Arthritis UK.

**Research in context:** *Evidence before this study:* Adverse childhood experiences are associated with increased risk of chronic pain, multimorbidity and premature mortality. DNA methylation-based epigenetic clocks are emerging biomarkers of biological aging and predictors of morbidity. Several studies have examined associations between childhood adversity and epigenetic age acceleration, but findings remain heterogeneous across cohorts, adversity measures and clock types. Chronic pain has also been linked to altered biological aging profiles. However, no previous study has integrated childhood trauma, biological aging and chronic pain within a unified analytical framework across multiple generations of epigenetic clocks. In addition, few studies have stratified by sex despite evidence that women exhibit greater chronic pain burden and multimorbidity across the lifespan.

*Added value of this study:* Using two independent population cohorts from the UK and USA, we demonstrate that childhood trauma is associated with both chronic pain and epigenetic age acceleration, with associations preferentially captured by second- and third-generation epigenetic clocks. These relationships were more consistent in women than men and were dose-dependent with trauma severity. Furthermore, epigenetic age acceleration established using GrimAge clocks, partially mediates the relationship between childhood trauma and chronic pain in women. Sequential adjustment models and sensitivity analyses additionally demonstrated that smoking-related biological aging pathways contribute substantially to these relationships, particularly in women.

*Implications of all the available evidence:* These findings support the hypothesis that childhood trauma contributes to chronic pain vulnerability partly through accelerated biological aging. The preferential involvement of second-generation epigenetic clocks suggests that morbidity-related aging measures capture the long-term biological consequences of early-life adversity more sensitively than chronological age predictors. The stratified analyses indicate that biological embedding of childhood trauma likely differs between women and men and may contribute to sex-specific trajectories of morbidity. Additional studies are required to determine whether accelerated biological aging following childhood trauma is modifiable through behavioural, clinical, or public health interventions.

## Introduction

Childhood trauma, encompassing emotional, physical, and sexual abuse, and emotional and physical neglect, is associated with enduring consequences for adult health. These include poor mental health, substance misuse, cardiometabolic disease, multimorbidity, premature mortality, and chronic pain.^1–4^ Chronic pain is particularly relevant to healthy longevity because it is common, disabling, frequently co-occurs with other long-term conditions and is associated with reduced function and quality of life.^5^ However, the biological processes through which childhood trauma increases later-life vulnerability to pain and other poor health outcomes remain poorly understood.

Epigenetic processes, including DNA methylation, are mechanisms by which exposure to severe or persistent stress can become ingrained.^6,7^ DNA methylation is particularly relevant because it has been linked to aging, morbidity, and mortality. Most biological processes are affected by aging, and many biomarkers have been used to generate machine learning models (biological clocks) that estimate age from features that vary across the lifespan.^8^ Biological ageing can be quantified using epigenetic clocks that use distinctive patterns of DNA methylation. First-generation clocks, including Horvath and Hannum, were developed primarily to predict chronological age.^9,10^ By contrast, second-generation clocks were trained to capture morbidity. The PhenoAge clock incorporates DNA methylation surrogates of physiological dysregulation, whereas GrimAge incorporates methylation surrogates of smoking exposure and circulating proteins associated with mortality.^11–15^ DunedinPACE estimates the pace of biological aging rather than attained biological age.^14^ These differences in approach are potentially important because childhood trauma may be more strongly associated with aging measures that capture aspects of health deterioration than with clocks optimised to estimate chronological age.

Our recent systematic review and meta-analysis found heterogeneous evidence linking adverse childhood experiences to DNA methylation-derived age acceleration.^6^ Interpretation of the literature was limited by inconsistent assessment of adversity, variable covariate adjustment and limited replication in independent cohorts. Furthermore, few studies tested whether biological aging is related to clinically important consequences of childhood trauma. Chronic pain is an important outcome in this context because it is linked to childhood adversity and to inflammatory, metabolic, neuroendocrine and behavioural pathways also implicated in biological aging. Emerging evidence suggests altered biological aging profiles in individuals with pain, but whether epigenetic age acceleration links childhood trauma with chronic pain remains unknown.^16,17^

Importantly, sex may modify the relationships between childhood trauma, biological ageing, and chronic pain. Women have a higher burden of chronic pain and multimorbidity than men.^5,18^ Sex-stratified analyses are therefore important when examining childhood trauma, biological aging, and pain.

This study, as part of the Consortium Against Pain Inequality (CAPE), investigated associations between childhood trauma, chronic pain, and DNA methylation-derived biological aging in Generation Scotland (GS) and Midlife in the United States (MIDUS). We examined multiple epigenetic clocks spanning three generations and evaluated whether biological aging mediated associations between childhood trauma and chronic pain in women and men.

## Methods

### Study design and participants

This analysis followed our systematic review and meta-analysis and a preregistered plan.^6,19^ We analysed two population cohorts with available childhood trauma, DNA methylation and chronic pain data. GS: Scottish Family Health Study is a longitudinal Scottish cohort recruited from 2006 onwards.^20^ Childhood trauma data were available from the STratifying Resilience and Depression Longitudinally (STRADL) sub-study, and blood DNA methylation data were derived from baseline samples collected between 2006 and 2011.^21,22^ MIDUS is a longitudinal US study established in 1995– 96, with blood collected during the MIDUS 2 biomarker assessment approximately 10 years later.^23,24^ Participants were included if they had complete childhood trauma, DNA methylation, and chronic pain data, providing analytical samples of 901 participants in GS and 1041 in MIDUS (Table 1).

**Table 1.**
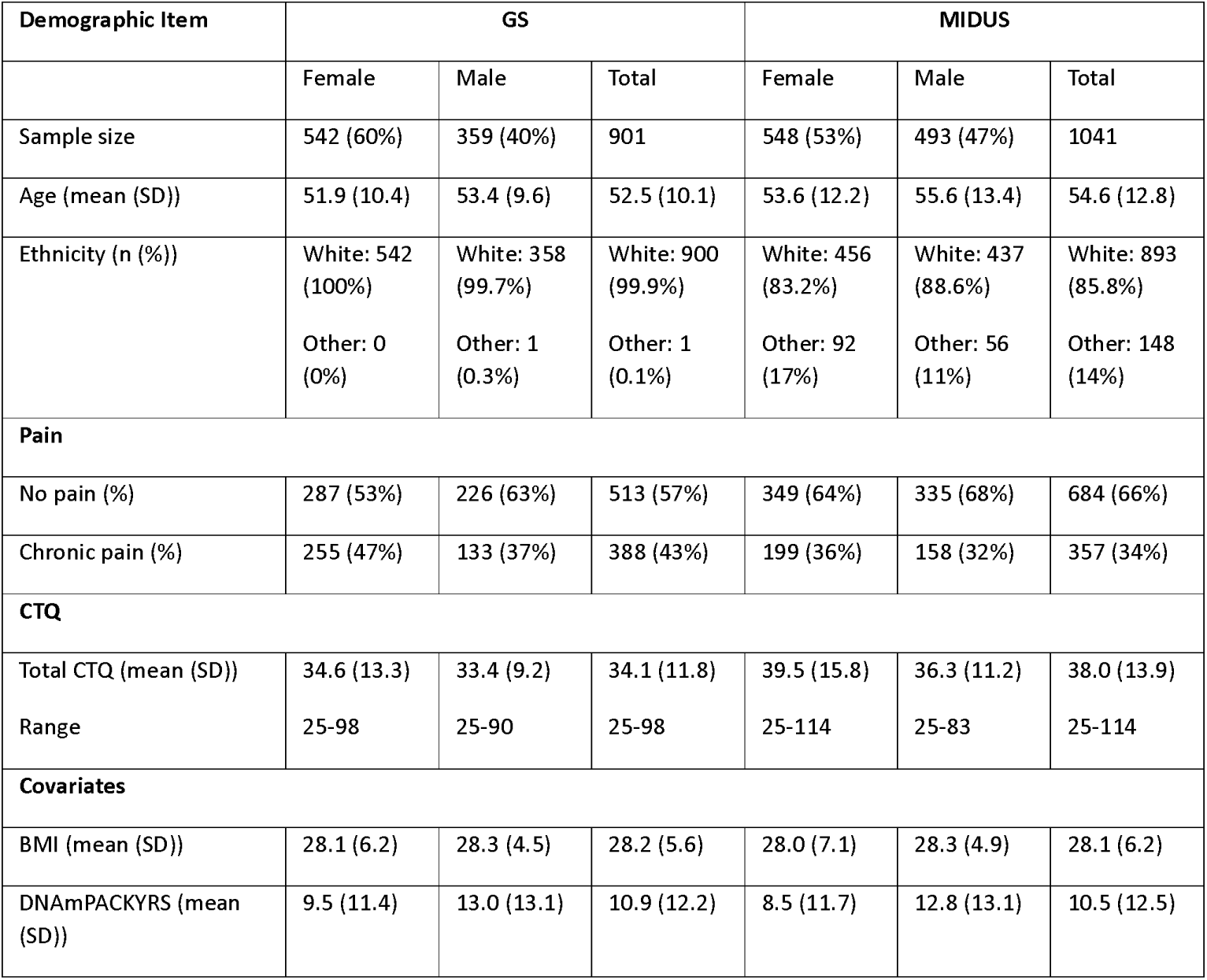
Cohort demographics.

### Childhood trauma and chronic pain

Childhood trauma was assessed retrospectively using the Childhood Trauma Questionnaire (CTQ).^25^ The CTQ measures emotional, physical, and sexual abuse and emotional and physical neglect. Subscale scores range from 5 to 25 and total scores from 25 to 125, with higher scores indicating greater exposure. Subscale and total CTQ scores were analysed continuously. For severity exposure– response analyses, total CTQ scores were categorised using established CTQ total-score severity thresholds: none/minimal (≤36), low (37–51), moderate (52–68) and severe (≥69).

In GS, chronic pain was defined as current continuous or intermittent pain lasting at least 3 months. In MIDUS, participants were classified as having chronic pain if they reported pain persisting beyond normal healing for several months or longer.

### DNA methylation processing and biological age measures

DNA methylation was measured (both cohorts) in blood using the Illumina Infinium MethylationEPIC BeadChip v1 and raw intensity files were processed consistently across both cohorts using R version 4.5.0 with minfi.^26^ Samples were removed if at least 10% of probes had detection p values greater than 0.05 or fewer than three beads (Figure S1). Probes were excluded if they failed equivalent quality thresholds in at least 10% of samples. Sex chromosome probes, cross-reactive probes and probes containing or overlapping common polymorphisms were also removed. Data were standardised using normal-exponential out-of-band (noob) normalisation. Batch effects were adjusted on M-values using ComBat, excluding batches containing fewer than three samples. Corrected M-values were converted to β-values for age estimation.

DNA methylation ages for Horvath, Hannum, PhenoAge, GrimAge and GrimAge2 were calculated using the Clock Foundation DNA Methylation Age Calculator. DunedinPACE was calculated as a rate measure using its R package.^14^ Epigenetic age acceleration (EAA) for age-estimating clocks was defined as the residual from regression of DNA methylation age on chronological age.

### Covariates and analytical strategy

Analyses of EAA were adjusted for blood-cell proportions, estimated by the DNA methylation age calculator (Figure S2). Estimated proportions of granulocytes, monocytes, CD4+ T cells, CD8+ T cells, B cells and natural killer cells were included as a confounder in all analyses of EAA. In addition, the causal framework identified ethnicity and childhood socioeconomic status (SES) as potential pre-exposure confounders and analyses were stratified by sex (Figure S3). Participant’s educational attainment, harmonised across cohorts as low, medium, or high, was used as a proxy for childhood SES. Relationships between CTQ score and participant educational attainment (Figure S4) were similar to those observed using parental educational attainment in MIDUS, supporting its use as a harmonised SES proxy while recognising the possibility of residual over-adjustment (Figure S5). BMI and smoking were treated as potential downstream covariates rather than primary confounders (Figure S3). BMI was log-transformed, and smoking was quantified by DNAmPackYrs, a DNA methylation-derived estimate of smoking history, and Yeo-Johnson transformed.^13^

Primary analyses were performed separately in GS and MIDUS and combined (pooled) by meta-analytic synthesis in accordance with the preregistered plan.^19^ Three sequential sex-stratified confounder/covariate adjustment models were used: Model 1 adjusted for ethnicity and SES. Chronological age was also adjusted for explicitly when the analysis involved DunedinPACE and in all chronic pain analyses (EAA analyses involving other clocks already included chronological age due to the EAA residual being used); Model 2 also included BMI; and Model 3 additionally included DNAmPackYrs. Associations between CTQ scores and chronic pain were examined using logistic regression. Associations between CTQ scores and EAA were estimated using linear regression. Estimates are presented as standardised β coefficients or odds ratios with 95% confidence intervals. Secondary analyses of severity-dependent associations used combined datasets.

Mediation analyses in sequentially adjusted sex stratified models assessed whether EAA mediated associations between total CTQ score and chronic pain. For each clock, linear models estimated associations between CTQ and EAA and logistic models estimated chronic pain from CTQ and EAA. Average causal mediation effects were estimated using 1000 non-parametric bootstrap simulations.

### Ethics and governance

Generation Scotland received ethics approval from the NHS Tayside Committee on Medical Research Ethics (05/S1401/89). MIDUS was approved by the relevant institutional review boards, including the University of Wisconsin Madison. Participants provided written informed consent. Data were accessed through approved governance procedures and analysed in de-identified form in a secure research environment.

## Results

### Participant characteristics

The analytical samples comprised 901 participants from Generation Scotland (GS) and 1041 participants from MIDUS with complete childhood trauma, DNA methylation and chronic pain data (Table 1). Chronic pain was more prevalent in women than men in both cohorts and women had higher upper-range CTQ scores across several trauma domains (Table S1).

### Childhood trauma is associated with chronic pain

Higher total CTQ score was associated with increased odds of chronic pain in GS and MIDUS and in pooled analyses (Figure 1A; Table S2). In the confounder-adjusted Model 1, associations were significant in women (OR 1·57, 1·37-1·81, per SD increase in CTQ) and in men (OR 1·27, 1·10-1·47) and were directionally consistent across cohorts.

**Figure 1.**
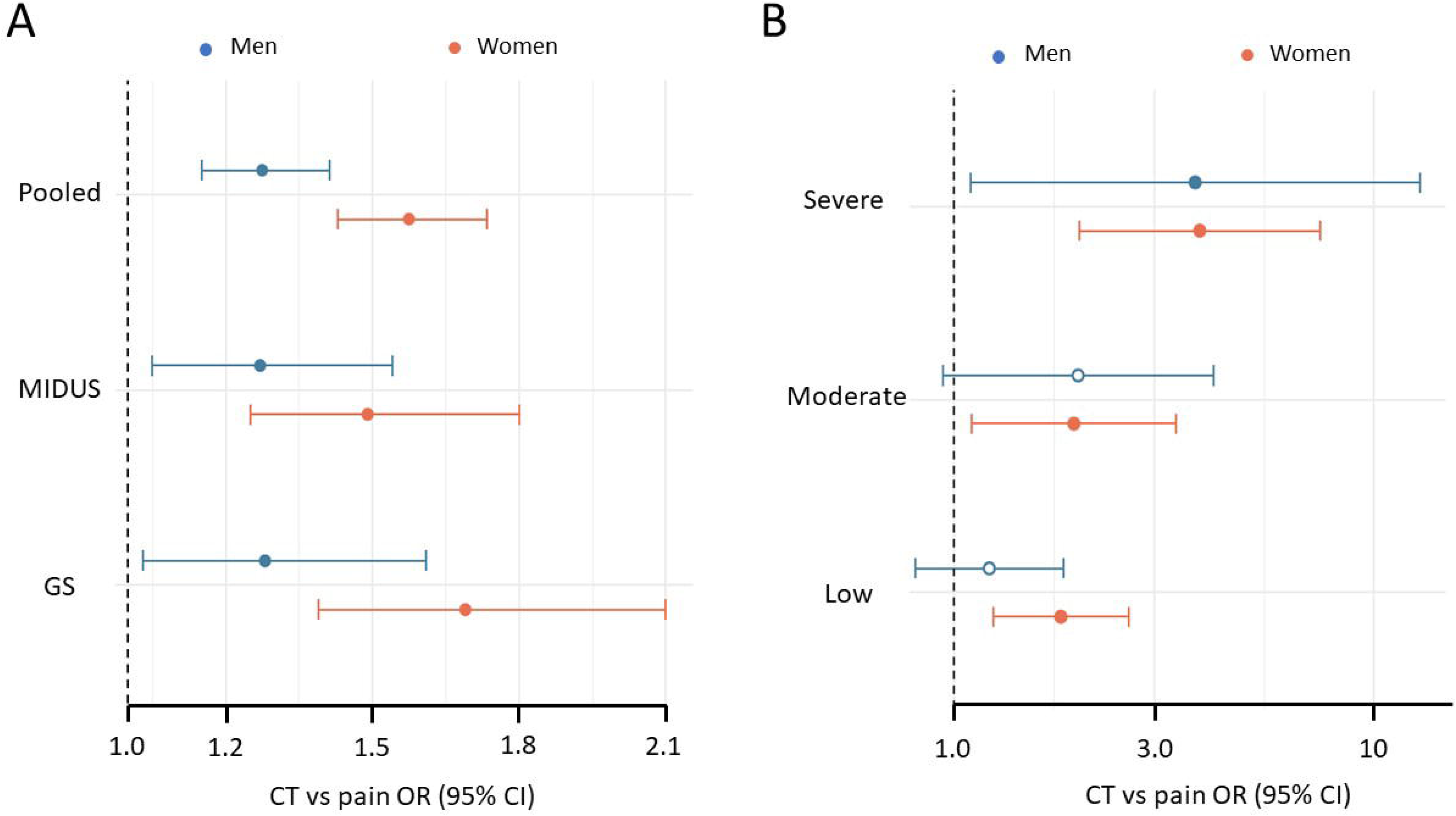
Associations between childhood trauma and chronic pain. A, Associations between childhood trauma and pain were significant in women and men of GS and MIDUS, and in the pooled meta-analysis. B, In a combined analysis across both cohorts the strength of the association depends on the severity of exposure to childhood trauma.

The confounder-adjusted (Model 1) associations were only modestly attenuated by sequential covariate adjustment. Following full adjustment for both BMI and DNAmPackYrs (Model 3), the pooled association between total CTQ score and chronic pain remained in women (OR 1·47, 1·27-1·69) and in men (OR 1·26, 1·09-1·45). Similar patterns were observed across individual trauma domains (Table S2).

In the combined confounder adjusted analysis, associations were graded by trauma severity (Figure 1B; Table S3). Severe childhood trauma was associated with substantially higher odds of chronic pain (OR 3·76, 95% CI 2·11-6·69) and this association remained after adjustment (Model 3) for BMI and DNAmPackYrs, the DNA methylation smoking proxy (OR 3·15, 1·75-5·66). The severity-dependent pattern was apparent in women and men.

### Childhood trauma is associated with increased biological aging

In confounder-adjusted analyses, childhood trauma was associated with biological aging measured using second-generation clocks and DunedinPACE, but not with first-generation clocks (Tables S4). For GrimAge clocks, the association survived sequential adjustment and was consistently significant across cohorts in women but not in men. In GrimAge (Figure 2A), for the pooled analysis, β is 0·161 (0·105-0·217) for women compared to a β of 0·071 (0·008-0·134) for men. Adjustment for BMI (Model 2) had relatively little effect (Table S4). However, the strength of the association between trauma and EAA was substantially diminished in Model 3, with inclusion of adjustment for DNAmPackYrs. In women, the pooled association between CTQ score and GrimAge acceleration decreased from β 0·161 (95% CI 0·105-0·217) in the confounder-adjusted model to 0·147 (0·090-0·203) after BMI adjustment and 0·050 (0·014-0·087) after additional adjustment for DNAmPackYrs. A similar pattern was observed for GrimAge2 and DunedinPACE. Associations between trauma and EAA for PhenoAge were small and Horvath and Hannum measures showed no consistent association with childhood trauma across models (Table S4).

**Figure 2.**
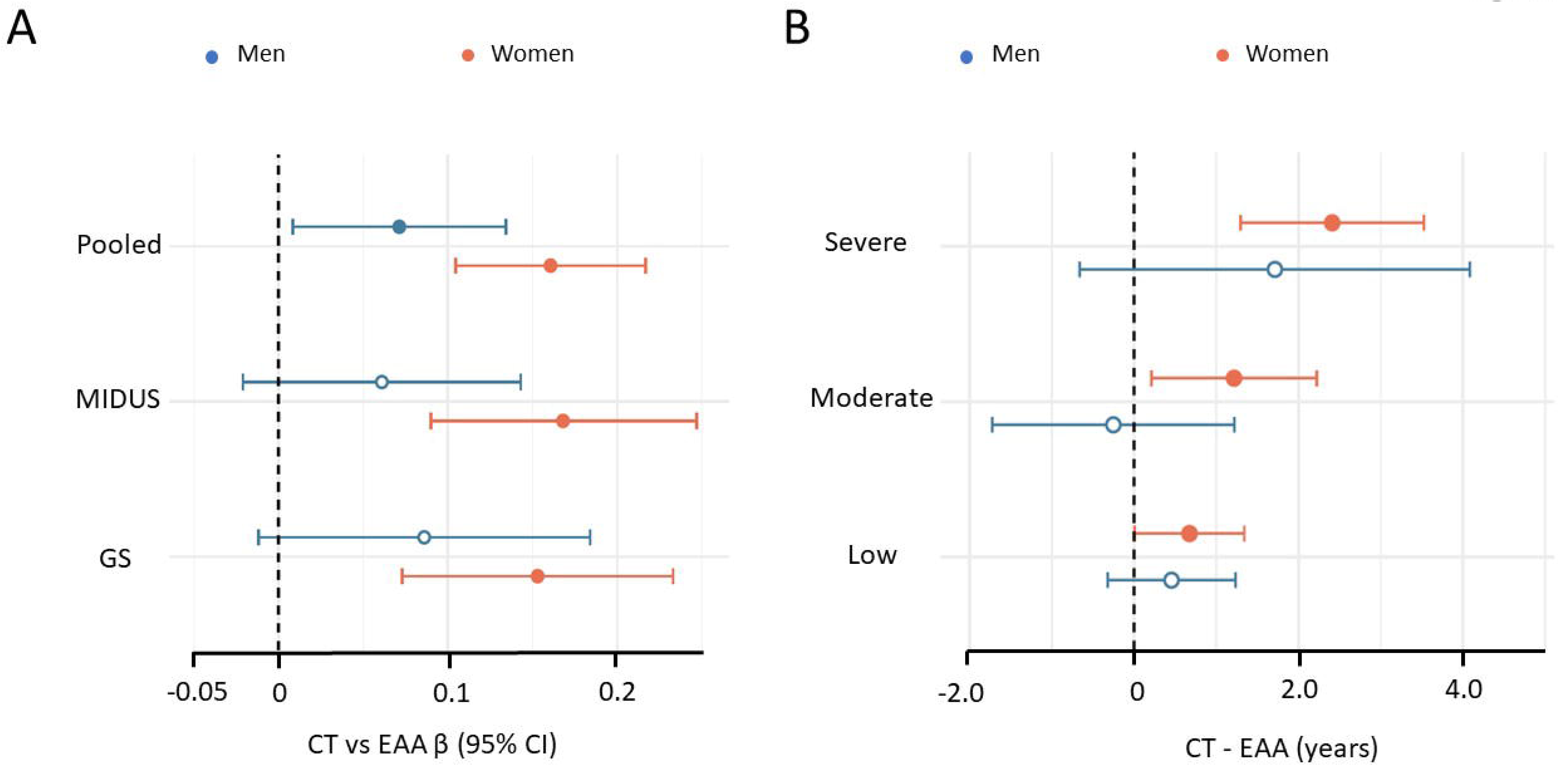
Associations between childhood trauma and epigenetic age acceleration. A, Regression analysis demonstrates significant associations between childhood trauma and EAA for women, and in pooled analysis for men and women, using the GrimAge clock. B, In the combined dataset the association is severity dependent.

Increasing trauma severity was associated with progressively greater GrimAge acceleration, particularly in women (Figure 2B) a relationship shared by GrimAge2, PhenoAge and DunedinPACE, but not Horvath or Hannum (Table S5). Among women, severe trauma was associated with 2·41 years of GrimAge acceleration in the confounder-adjusted model (95% CI 1·29-3·53), remaining after BMI adjustment (2·23 years, 1·11-3·35), but reduced following additional adjustment for DNAmPackYrs (0·87 years, 0·14-1·60). These findings indicate that childhood trauma-related differences in morbidity-linked biological aging overlap substantially with smoking-related methylation signatures.

### Associations between biological aging and chronic pain involve smoking

Associations between biological aging and chronic pain were also most consistent in GrimAge measures and in women (Table S6). In pooled confounder-adjusted analyses of women, higher GrimAge acceleration was associated with increased odds of chronic pain (OR 1·29, 95% CI 1·13-1·48, per SD increase in EAA), with little change after adjustment for BMI (OR 1·28, 1·11-1·47). Following additional adjustment for DNAmPackYrs, the positive association was no longer evident (OR 0·87, 0·70-1·09). GrimAge2 showed the same pattern, while no significant corresponding associations were observed in men. EAA calculated using the other clocks showed no consistent associations with chronic pain for women or men.

### GrimAge partially mediated associations between childhood trauma and chronic pain in women

Age acceleration established by GrimAge clocks alone partially mediated the association between childhood trauma and chronic pain in women (Table S6), with mediation effects observed in MIDUS and in the pooled analyses. In confounder adjusted (Model 1) pooled analyses (Figure 3), the average causal mediation effect for GrimAge in women was modest (0·0068, 0·001-0·012). GrimAge2 EAA had a similar mediation effect in women (Table S7), however, corresponding effects were not observed in men. Furthermore, no other epigenetic clock significantly mediated associations between childhood trauma and chronic pain in women or men.

**Figure 3.**
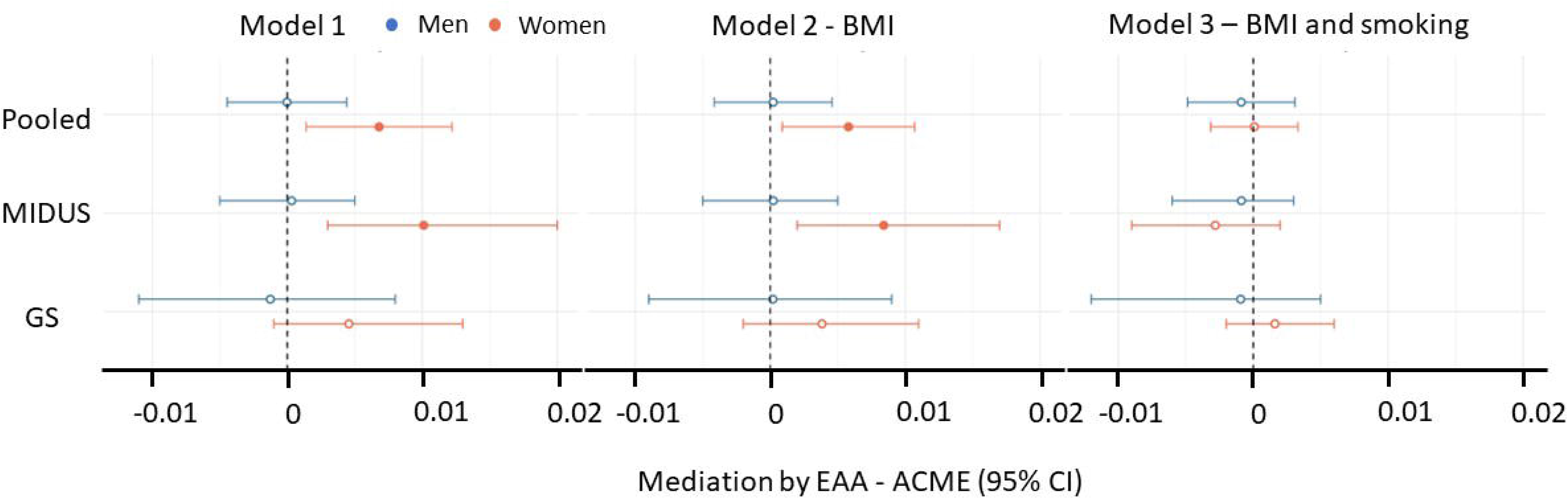
Mediation of the association between childhood trauma and chronic pain by GrimAge acceleration. Mediation effect is abolished by adjustment for smoking, but not BMI.

The mediation effect of GrimAge in women remained evident in Model 2 after adjustment for BMI (0·0058, 0·001–0·011) but was absent in Model 3 after additional adjustment for DNAmPackYrs (Figure 3). Similar attenuation after sequential adjustment was also observed for GrimAge2 (Table S7).

Together, these findings suggest that GrimAge morbidity-related aging partially accounts for the association between childhood trauma and chronic pain in women, with smoking contributing substantially to this relationship.

## Discussion

In two independent population cohorts, childhood trauma was associated with chronic pain and accelerated biological aging, particularly when aging was measured using morbidity-related clocks. These relationships were generally stronger in women than men and increased with trauma severity. In women, GrimAge acceleration statistically mediated a modest component of the relationship between childhood trauma and chronic pain, an effect that was abolished after adjustment for DNA methylation markers of smoking (DNAmPackYrs). These findings suggest that biological aging signatures related to later-life health risk may reflect the long-term embodiment of childhood trauma. The relationships between childhood trauma, biological aging and chronic pain are particularly strong in women and substantially involve smoking.

The association between childhood trauma and chronic pain is consistent with previous population-based evidence linking adverse childhood experiences with pain and multimorbidity in adulthood.^1–4^ Our findings extend this literature by showing a severity-dependent relationship between childhood trauma and chronic pain across two cohorts located in different continents and by demonstrating that the association remained after consideration of confounders and systematic adjustment for covariates.

The study also significantly advances the existing literature by demonstrating the specificity of childhood trauma associations with biological aging according to the epigenetic measure used.^6^ Childhood trauma was associated with age acceleration using GrimAge, PhenoAge and DunedinPACE, whereas associations with Horvath and Hannum clocks were weak or absent. This is consistent with the distinction between clocks trained to estimate chronological age and those designed to reflect morbidity, or pace of aging.^9–15^ Childhood trauma was not uniformly related to all biological aging measures. The clearest associations were observed for clocks designed to capture morbidity, or pace of aging. This distinction is clinically important because such signatures may help to describe the processes through which early-life adversity is associated with reduced healthy longevity.

GrimAge clock outputs were particularly informative. Childhood trauma severity was associated with progressively greater GrimAge acceleration, especially in women. The sequentially adjusted models showed marked attenuation after inclusion of DNAmPackYrs which indicates that the observed association in GrimAge substantially involves lifetime smoking load. Smoking likely represents a downstream behavioural pathway associated with childhood trauma that contributes to biological aging. However, smoking-related DNA methylation signatures are also intrinsic components of GrimAge algorithms and adjustment for DNAmPackYrs may partially remove biologically relevant variance captured by these clocks. The importance of smoking was also apparent in analyses linking biological aging with chronic pain. In women, GrimAge acceleration was positively associated with chronic pain in confounder-adjusted models and remained so after adjustment for BMI with 20% higher odds of chronic pain for each standard deviation increase in GrimAge acceleration. The association was abolished after additional adjustment for DNAmPackYrs. Furthermore, GrimAge clocks statistically mediated a modest component (approximately 10-15%) of the association between childhood trauma and chronic pain in women, whereas no mediation was evident in men or in either sex for the other biological aging measures. These findings suggest that smoking-related exposure represents a pathway through which childhood trauma is associated with later-life pain vulnerability in women. The greater magnitude and consistency of the relationships in women are relevant to healthy longevity because women experience the highest burden of chronic pain and multimorbidity across adulthood.^5,18^ Childhood trauma may therefore contribute to sex-specific trajectories of pain-related ill health across the lifespan.

Biological aging likely represents only one component of a broader network of pathways linking childhood trauma to chronic pain. Previous work within CAPE demonstrated that mental ill-health accounts for a substantial proportion of the association between childhood adversity and chronic widespread pain.^4^ Childhood trauma may therefore influence pain vulnerability through interacting behavioural and biological aging mechanisms rather than through a single dominant pathway.

Women had higher upper-range CTQ scores, more chronic pain and stronger associations of childhood trauma with morbidity-related biological aging. This pattern is consistent with population evidence that women experience a higher burden of several forms of childhood adversity and may show greater long-term health consequences of adversity exposure.^2,27^ In the context of pain, nociceptive processing, stress responsivity, affective symptoms, and vulnerability to trauma- and stress-related disorders may contribute to the stronger associations observed here.^28,29^ These mechanisms cannot be resolved by the present analyses, but they support the importance of sex-stratified approaches when examining biological embedding of childhood trauma.

This cross-sectional study has some limitations. First, childhood trauma was retrospectively assessed, and DNA methylation was also measured in adulthood. While the causal sequence cannot be established here, previous evidence for biological age acceleration in children exposed to maltreatment suggest that this likely occurs early and is therefore a plausible mediator of poor health outcomes throughout life.^30^ Second, chronic pain was defined differently in GS and MIDUS. Although the consistency of findings across cohorts supports robustness, the measures do not capture pain severity, impact, or spread, factors that may strengthen relationships with childhood trauma.^4^ Consequently, the observed associations likely underestimate relationships with more severe or disabling pain phenotypes. Third, educational attainment was used as a proxy for childhood socioeconomic status across cohorts but may itself be influenced by trauma exposure and therefore treating it as a confound may cause over-adjustment. However, similar relationships were seen in MIDUS using parental education as an alternative proxy that is upstream of participants’ childhood exposures. Finally, the GS cohort is a predominantly white population, and ethnic diversity is restricted in MIDUS, which potentially limits generalisability across global populations.

Strengths include replication of associations across independent UK and US cohorts, harmonised DNA methylation processing, analysis of multiple clock generations, sex-stratified and trauma severity analyses and a prespecified analytical approach guided by a systematic review and meta-analyses.^6, 19^ Importantly, the comparison of models before and after smoking adjustment enabled interpretation of the substantial overlap between childhood trauma-related aging signatures, smoking-related DNA methylation and chronic pain.

In conclusion, childhood trauma was associated with chronic pain and with biological aging signatures related to morbidity, particularly in women. GrimAge acceleration accounted for a small component of the association between childhood trauma and pain in women, with findings implicating smoking-related methylation pathways. Our results support the hypothesis that early-life adversity contributes to diminished healthy longevity through interacting behavioural and biological processes with chronic pain as an important later-life manifestation of this vulnerability.

## Data Availability

All data produced in the present study are available upon reasonable request to the corresponding author.

## Author contributions

Study design, conceptualisation, methodology and data interpretation: HR, SS, GJM, SC, CGB, TGH; data analysis, visualisation: HR, SS, TGH; writing, review, and editing: HR, SS, LAC, GJM, SC, CGB, TGH; funding acquisition: LAC, GJM, TGH; supervision, project administration: TGH.

## Data availability

Data will be made available upon reasonable request.

## AI declaration statement

ChatGPT (GPT-5.5 Thinking, OpenAI) was used during preparation of this manuscript, to assist with language editing and refinement of the draft text. The tool was not used to generate or analyse data. All content was reviewed, edited, and verified by the authors, who take full responsibility for the accuracy, integrity, and final content of the manuscript.

## Acknowledgements

The HR, SS, LAC, GJM and TGH are members of the Advanced Pain Discovery Platform and the Consortium against Pain Inequality (CAPE) studying the impact of adverse childhood experiences on pain. Members of the CAPE advisory group, through their lived experiences, have guided the project, including recognition of the need for evidence of severity-dependant impacts of childhood trauma and biomarkers of vulnerability to chronic pain.

## References

1 Felitti VJ, Anda RF, Nordenberg D, Williamson DF, Spitz AM, Edwards V, Koss MP, Marks JS. Relationship of childhood abuse and household dysfunction to many of the leading causes of death in adults: the Adverse Childhood Experiences (ACE) Study. Am J Prev Med. 1998; 14(4): 245– 258.

2 Hughes K, Bellis MA, Hardcastle KA, Sethi D, Butchart A, Mikton C, Jones L, Dunne MP. The effect of multiple adverse childhood experiences on health: a systematic review and meta-analysis. Lancet Public Health. 2017; 2(8): e356–e366.

3 Senaratne DNS, Thakkar B, Smith BH, Hales TG, Marryat L, Colvin LA. The impact of adverse childhood experiences on multimorbidity: a systematic review and meta-analysis. BMC Med. 2024; 22(1): 315.

4 Timmins KA, Hales TG, Macfarlane GJ; Consortium Against Pain InEquality (CAPE) investigators and Chronic Pain Advisory Group. Childhood maltreatment and chronic "all over" body pain in adulthood: a counterfactual analysis using UK Biobank. Pain. 2025; 166(5): 1204–1211.

5 Rometsch C, Martin A, Junne F, Cosci F. Chronic pain in European adult populations: a systematic review of prevalence and associated clinical features. Pain. 2025; 166(4): 719–731.

6 Russell H, Angus G, Singleton S, Bell CG, Hales TG. The impact of adverse childhood experiences on DNA methylation age: a systematic review and meta-analysis. Clin Epigenetics. 2026; 18(1): 33.

7 Zhou A, Ryan J. Biological Embedding of Early-Life Adversity and a Scoping Review of the Evidence for Intergenerational Epigenetic Transmission of Stress and Trauma in Humans. Genes. 2023; 14(8): 1639.

8 Rutledge J, Oh H, Wyss-Coray T. Measuring biological age using omics data. Nat Rev Genet. 2022; 23(12): 715–727.

9 Horvath S. DNA methylation age of human tissues and cell types. Genome Biol. 2013; 14(10): R115.

10 Hannum G, Guinney J, Zhao L, Zhang L, Hughes G, Sadda S, Klotzle B, Bibikova M, Fan JB, Gao Y, Deconde R, Chen M, Rajapakse I, Friend S, Ideker T, Zhang K. Genome-wide methylation profiles reveal quantitative views of human aging rates. Mol Cell. 2013; 49(2): 359–367.

11 Marioni RE, Shah S, McRae AF, Chen BH, Colicino E, Harris SE, Gibson J, Henders AK, Redmond P, Cox SR, Pattie A, Corley J, Murphy L, Martin NG, Montgomery GW, Feinberg AP, Fallin MD, Multhaup ML, Jaffe AE, Joehanes R, Schwartz J, Just AC, Lunetta KL, Murabito JM, Starr JM, Horvath S, Baccarelli AA, Levy D, Visscher PM, Wray NR, Deary IJ. DNA methylation age of blood predicts all-cause mortality in later life. Genome Biol. 2015; 16: 25.

12 Levine ME, Lu AT, Quach A, Chen BH, Assimes TL, Bandinelli S, Hou L, Baccarelli AA, Stewart JD, Li Y, Whitsel EA, Wilson JG, Reiner AP, Aviv A, Lohman K, Liu Y, Ferrucci L, Horvath S. An epigenetic biomarker of aging for lifespan and healthspan. Aging. 2018; 10(4): 573–591.

13 Lu AT, Quach A, Wilson JG, Reiner AP, Aviv A, Raj K, Hou L, Baccarelli AA, Li Y, Stewart JD, Whitsel EA, Assimes TL, Ferrucci L, Horvath S. DNA methylation GrimAge strongly predicts lifespan and healthspan. Aging. 2019; 11(2): 303–327.

14 Belsky DW, Caspi A, Corcoran DL, Sugden K, Poulton R, Arseneault L, Baccarelli A, Chamarti K, Gao X, Hannon E, Harrington HL, Houts R, Kothari M, Kwon D, Mill J, Schwartz J, Vokonas P, Wang C, Williams BS, Moffitt TE. DunedinPACE, a DNA methylation biomarker of the pace of aging. eLife. 2022; 11: e73420.

15 Mavrommatis C, Belsky DW, Ying K, Moqri M, Campbell A, Richmond A, Gladyshev VN, Chandra T, McCartney DL, Marioni RE. An unbiased comparison of 14 epigenetic clocks in relation to 174 incident disease outcomes. Nat Commun. 2025; 16(1): 11164.

16 Cruz-Almeida Y, Fillingim RB, Riley JL 3rd, Woods AJ, Porges E, Cohen R, Cole J. Chronic pain is associated with a brain aging biomarker in community-dwelling older adults. Pain. 2019;160(5): 1119–1130.

17 Kwiatkowska KM, Bacalini MG, Sala C, Kaziyama H, de Andrade DC, Terlizzi R, Giannini G, Cevoli S, Pierangeli G, Cortelli P, Garagnani P, Pirazzini C. Analysis of Epigenetic Age Predictors in Pain-Related Conditions. Front. Public Health 2020; 8: 172.

18 Chowdhury SR, Chandra Das D, Sunna TC, Beyene J, Hossain A. Global and regional prevalence of multimorbidity in the adult population in community settings: a systematic review and meta-analysis. EClinicalMedicine. 2023; 57: 101860.

19 Russell H, Singleton S, Bell C, Macfarlane GJ, Cole S, Hales TG. The role of DNA methylation in the impact of adverse childhood experiences on chronic pain. OSF. 2025. https://osf.io/we76z

20 Smith BH, Campbell A, Linksted P, Fitzpatrick B, Jackson C, Kerr SM, Deary IJ, Macintyre DJ, Campbell H, McGilchrist M, Hocking LJ, Wisely L, Ford I, Lindsay RS, Morton R, Palmer CN, Dominiczak AF, Porteous DJ, Morris AD.. Cohort profile: Generation Scotland: Scottish Family Health Study (GS:SFHS). Int J Epidemiol. 2013; 42(3): 689–700.

21 Habota T, Sandu AL, Waiter GD, McNeil CJ, Steele JD, Macfarlane JA, Whalley HC, Valentine R, Younie D, Crouch N, Hawkins EL, Hirose Y, Romaniuk L, Milburn K, Buchan G, Coupar T, Stirling M, Jagpal B, MacLennan B, Priba L, Harris MA, Hafferty JD, Adams MJ, Campbell AI, MacIntyre DJ, Pattie A, Murphy L, Reynolds RM, Elliot R, Penton-Voak IS, Munafò MR, Evans KL, Seckl JR, Wardlaw JM, Lawrie SM, Haley CS, Porteous DJ, Deary IJ, Murray AD, McIntosh AM. Cohort profile for the STratifying Resilience and Depression Longitudinally (STRADL) study: A depression-focused investigation of Generation Scotland, using detailed clinical, cognitive, and neuroimaging assessments. Wellcome Open Res. 2021; 4: 185.

22 Seeboth A, McCartney DL, Wang Y, Hillary RF, Stevenson AJ, Walker RM, Campbell A, Evans KL, McIntosh AM, Hägg S, Deary IJ, Marioni RE. DNA methylation outlier burden, health, and ageing in Generation Scotland and the Lothian Birth Cohorts of 1921 and 1936. Clin Epigenetics. 2020; 12(1): 49.

23 Radler BT. The Midlife in the United States (MIDUS) series: a national longitudinal study of health and well-being. Open Health Data. 2014; 2(1): e3.

24 Love GD, Seeman TE, Weinstein M, Ryff CD. Bioindicators in the MIDUS national study: protocol, measures, sample, and comparative context. J Aging Health. 2010; 22(8): 1059–1080.

25 Bernstein DP, Stein JA, Newcomb MD, Walker E, Pogge D, Ahluvalia T, Stokes J, Handelsman L, Medrano M, Desmond D, Zule W. Development and validation of a brief screening version of the Childhood Trauma Questionnaire. Child Abuse Negl. 2003; 27(2): 169–190.

26 Zhuang BC, Jude MS, Konwar C, Yusupov N, Ryan CP, Engelbrecht HR, Whitehead J, Halberstam AA, MacIsaac JL, Dever K, Tran TK, Korinek K, Zimmer Z, Lee NR, McDade TW, Kuzawa CW, Huffman KM, Belsky DW, Binder EB, Czamara D, Korthauer K, Kobor MS. Accounting for differences between Infinium MethylationEPIC v2 and v1 in DNA methylation-based tools. Life Sci Alliance. 2025; 8(9): e202403155.

27 Stoltenborgh M, Bakermans-Kranenburg MJ, Alink LRA, van IJzendoorn MH. The prevalence of child maltreatment across the globe: review of a series of meta-analyses. Child Abuse Rev. 2015; 24(1): 37–50.

28 Fillingim RB, King CD, Ribeiro-Dasilva MC, Rahim-Williams B, Riley JL III. Sex, gender, and pain: a review of recent clinical and experimental findings. J Pain. 2009; 10(5): 447–485.

29 Li SH, Graham BM. Why are women so vulnerable to anxiety, trauma-related and stress-related disorders? The potential role of sex hormones. Lancet Psychiatry. 2017; 4(1): 73–82.

30 Chang OD, Meier HCS, Maguire-Jack K, Davis-Kean P, Mitchell C. Childhood Maltreatment and Longitudinal Epigenetic Aging: NIMHD Social Epigenomics Program. JAMA Netw Open. 2024; 7(7): e2421877.

