## Supplemental data for "Childhood trauma, accelerated biological aging and chronic pain across two population cohorts"

Figure S1: DNA methylation pre-processing and quality control

Figure S2: Associations between cell-type composition and EAA

Figure S3: Directed acyclic graph

Figure S4: Associations between childhood trauma and participant’s educational attainment

Figure S5: Associations in MIDUS between highest parental educational attainment and participant’s childhood trauma

Figure S6: Associations in MIDUS between ethnicity and CTQ, and ethnicity and EAA

Figure S7: Associations between CTQ and BMI and BMI and EAA

Figure S8: Associations between CTQ and smoking and smoking and EAA

Table S1: Childhood trauma categories

Table S2: Associations between childhood trauma and chronic pain

Table S3: Severity-dependent associations between childhood trauma and chronic pain

Table S4: Associations between childhood trauma and EAA

Table S5: Severity-dependent associations between childhood trauma and EAA

Table S6: Associations between EAA and chronic pain

Table S7: Associations between childhood trauma and pain - mediation by EAA

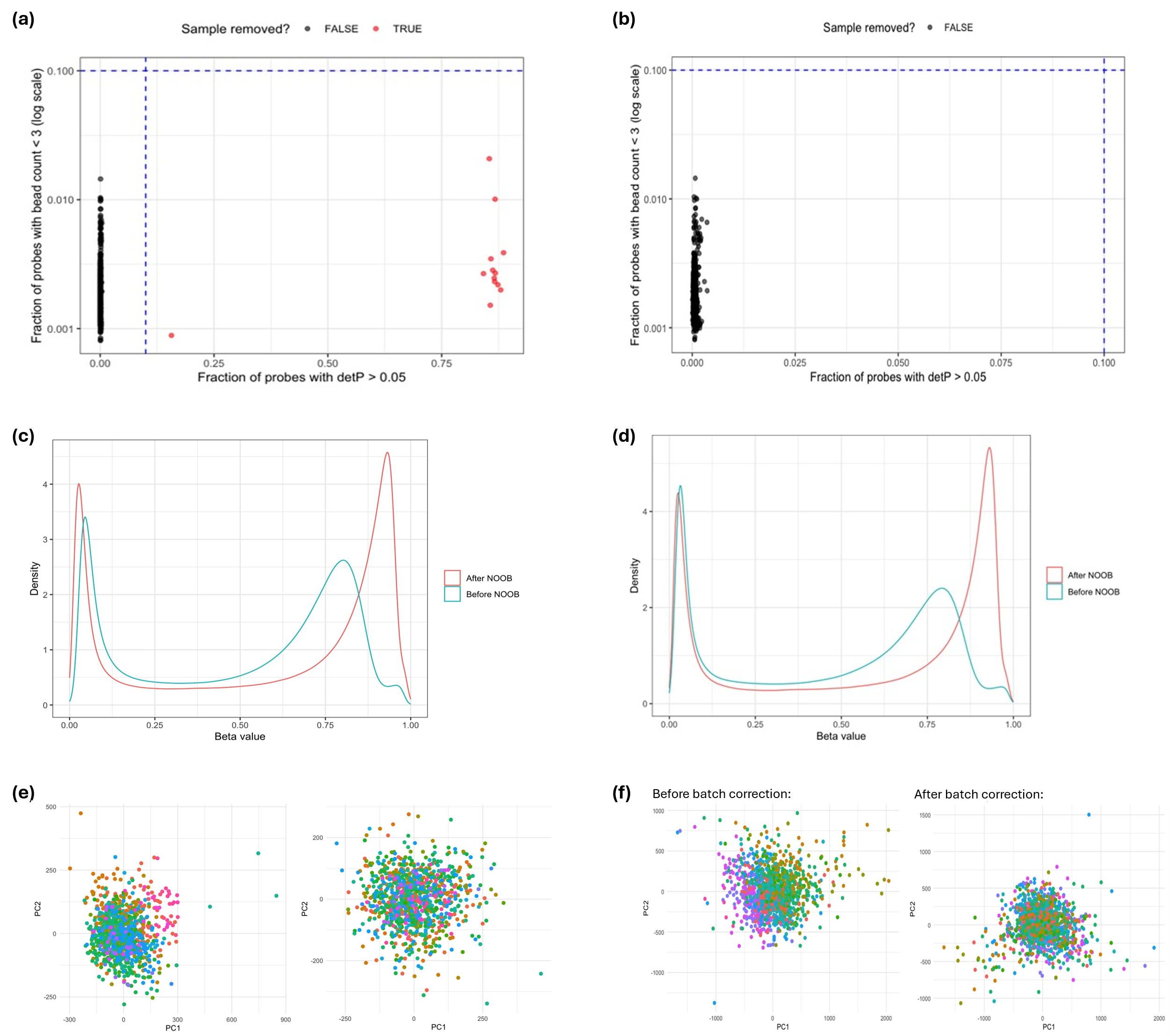

**Fig. S1.** DNA methylation pre-processing and quality control. GS (a,c,d) and MIDUS (b,d,f) data were subject to standard probe-level quality control methods, and (a,b) sample-level quality control before being (c,d) noob normalised and (e,f) corrected for batch effects (see Methods). Poor quality samples were identified and removed if detection p-values were > 0.05 in ≥ 10% probes, or if ≥ 10% probes possessed < 3 beads. Low quality probes were removed based on similar criteria. Probes were removed if detection p-values were > 0.05 in ≥ 10% samples, or if bead counts were < 3 in ≥ 10% samples. Probes located on the X and Y sex chromosomes were removed, alongside previously identified problematic probes, including those known to be cross-reactive, contain polymorphic CpGs, or overlap common SNPs (Zhou et al., 2017). Following quality control, data were normalised using the normal-exponential out-of-band (noob) method from the minfi package. M-values were calculated following normalisation for use in batch correction analysis. Samples were grouped according to processing batch and batches containing < 3 samples were excluded to ensure stability. Batch effects were corrected using ComBat facilitated by the sva package. Correction was performed on M-values, adjusting for batch while preserving biological variation. M-values were then converted to beta-values for use in epigenetic age calculations.

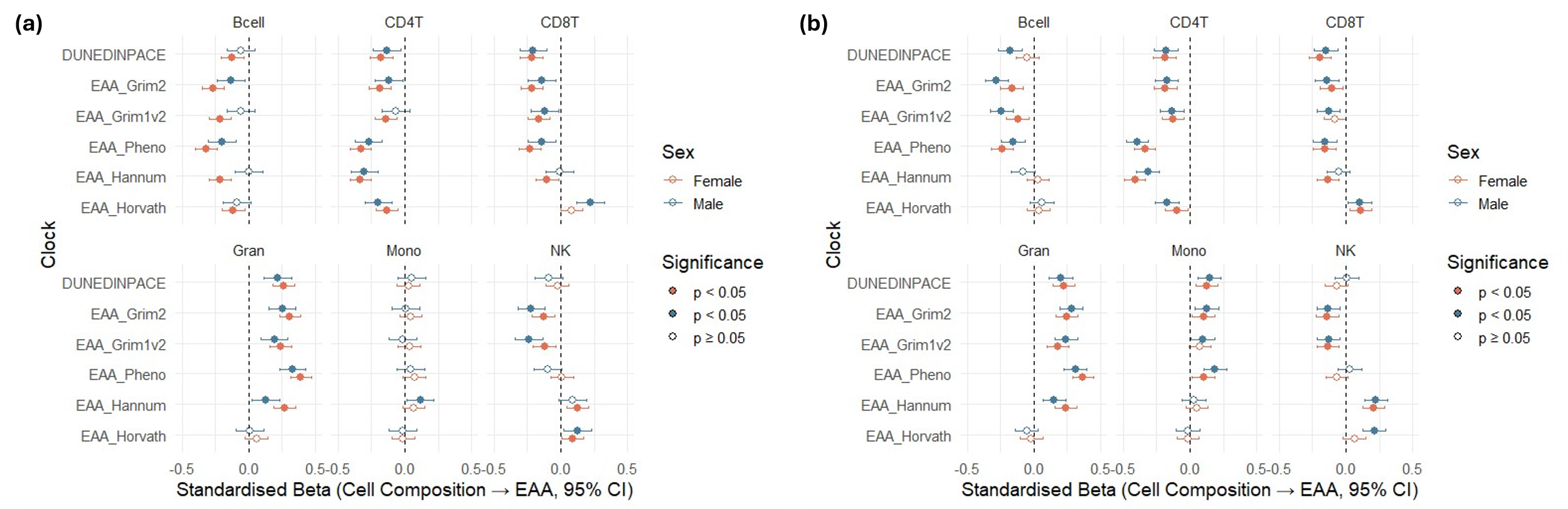

**Fig. S2.** Associations between cell-type composition and EAA. Analyses in (a) MIDUS and (b) GS were stratified by sex. The Clock Foundation output provides blood cell type composition estimates for granulocytes, monocytes, CD4+ T cells, CD8+ T cells, B cells and natural killer cells. EAA is epigenetic age acceleration, Bcell is B cells, CD4T is CD4+ T cells, CD8T is CD8+ T cells, Gran is granulocytes, Mono is monocytes and NK is natural killer cells. In general, second-generation clocks were more sensitive to cell composition than were first-generation clocks. Subsequent analyses were routinely adjusted for cell type composition. Variance inflation factor (VIF) tests were performed to assess potential multicollinearity. All VIFs were < 5 indicating no substantial multicollinearity.

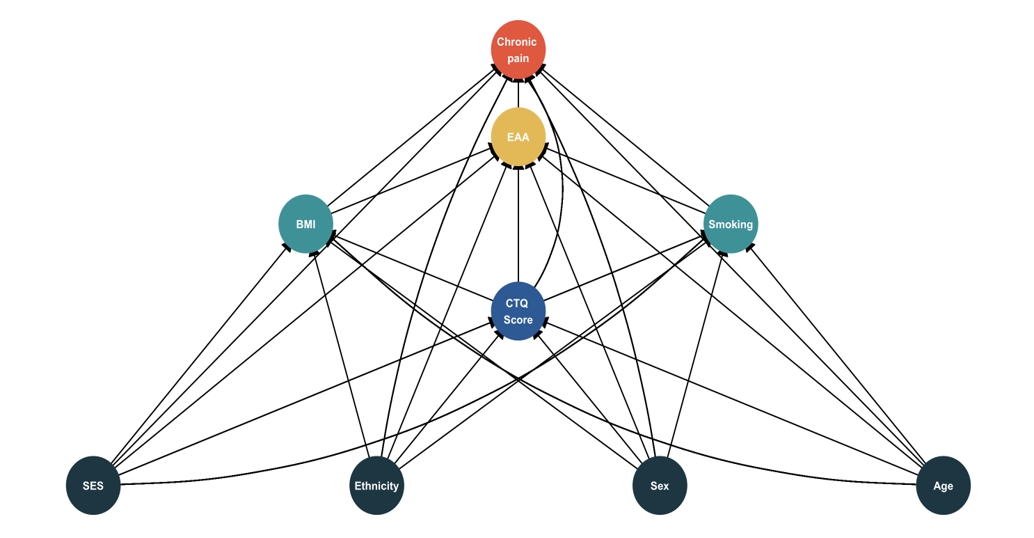

**Fig. S3.** Directed acyclic graph (DAG). Socioeconomic status (SES), ethnicity, sex, and age are potential confounders, while BMI and smoking are modelled as covariates.

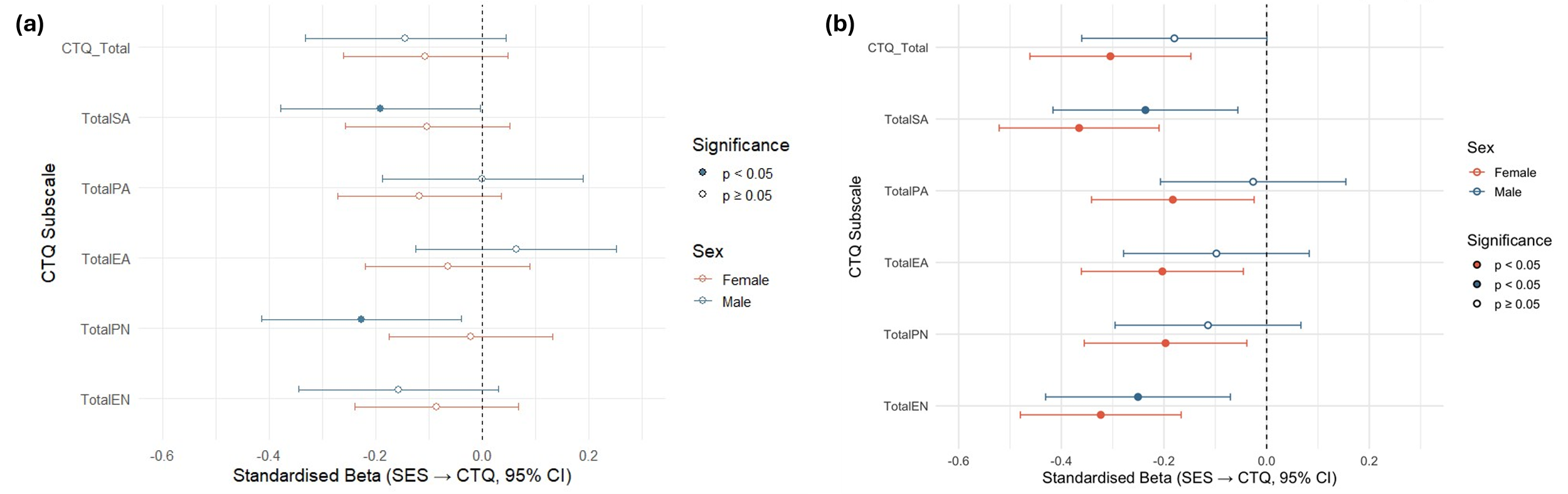

**Fig. S4.** Associations between participant’s educational attainment and childhood trauma. (a) GS and (b) MIDUS data were stratified by sex. CTQ Total is total CTQ score, TotalSA is total sexual abuse score, TotalPA is total physical abuse score, TotalEA is total emotional abuse score, TotalPN is total physical neglect score and Total EN is total emotional neglect score. SES is socioeconomic status, using the participant’s educational attainment as a proxy, and CTQ is childhood trauma questionnaire. In general, lower educational attainment by participants was associated with greater exposure to higher CTQ scores. Data were grouped into low, medium, and high educational attainment. Low indicates any level of education up to and including completion of high school, medium includes those participants who have completed UK college, US community college education, or any technical college/apprenticeship qualifications, or are currently studying for a higher education degree, while high indicates being awarded with a degree at bachelor level or above.

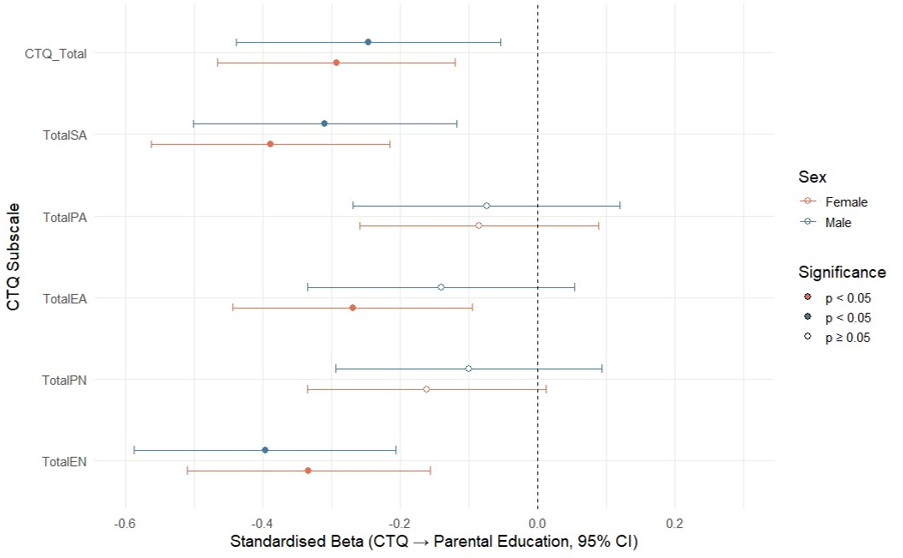

**Fig. S5.** Associations in MIDUS between highest parental educational attainment and participant’s childhood trauma. Parental educational attainment was not available in GS. Stratified by sex. CTQ_Total is total CTQ score, TotalSA is total sexual abuse score, TotalPA is total physical abuse score, TotalEA is total emotional abuse score, TotalPN is total physical neglect score and Total EN is total emotional neglect score. SES is socioeconomic position, using the parent’s highest educational attainment as a proxy, and CTQ is childhood trauma questionnaire. The tendency for higher CTQ scores in MIDUS participants whose parents had a lower educational attainment is very similar to the relationships seen in Figure S4. The same approach to categorisation of educational attainment described in Figure S4 was used here.

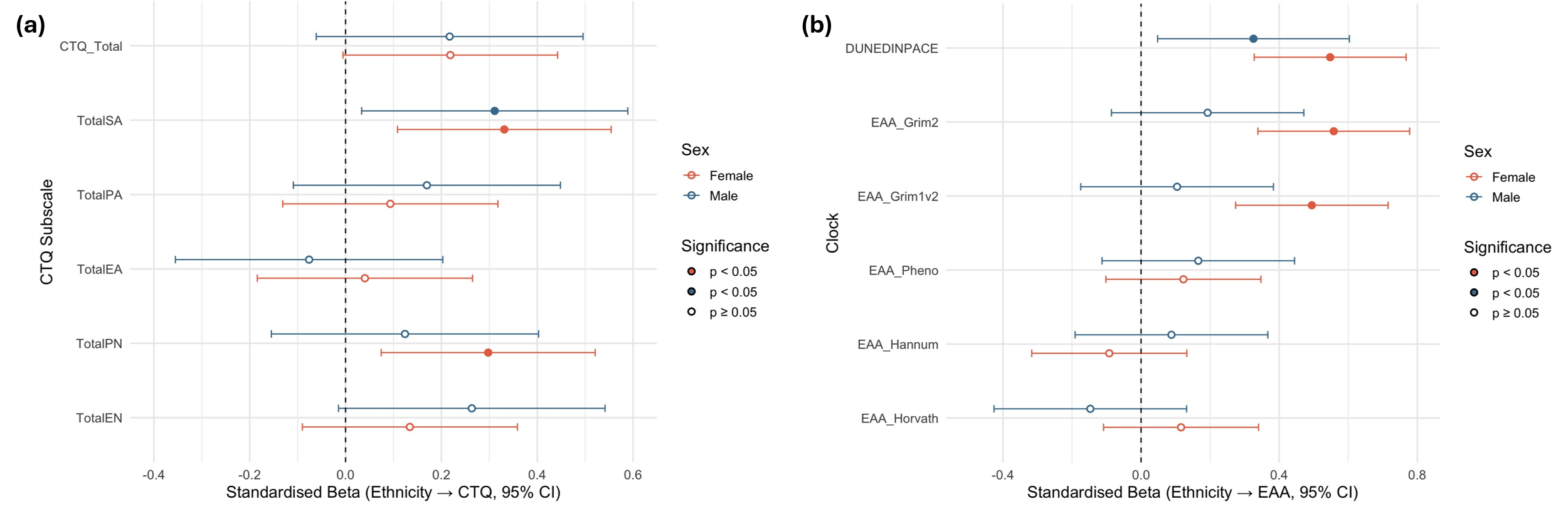

**Fig. S6.** Associations between ethnicity and CTQ score, and ethnicity and EAA in MIDUS. Data, ethnicity and CTQ score in (a) and ethnicity and EAA (b) were stratified by sex. CTQ_Total is total CTQ score, TotalSA is total sexual abuse score, TotalPA is total physical abuse score, TotalEA is total emotional abuse score, TotalPN is total physical neglect score and Total EN is total emotional neglect score. EAA is epigenetic age acceleration, and CTQ is childhood trauma questionnaire. In general, identifying as an ethnicity other than white was associated with higher CTQ scores and greater EAA. The association between other than white ethnicity and EAA were restricted to GrimAge and DundedinPACE measures and greatest for women. Participants were grouped into white or ‘other ethnic group’, as the numbers of participants identifying as other than white were relatively small making further stratification unfeasible.

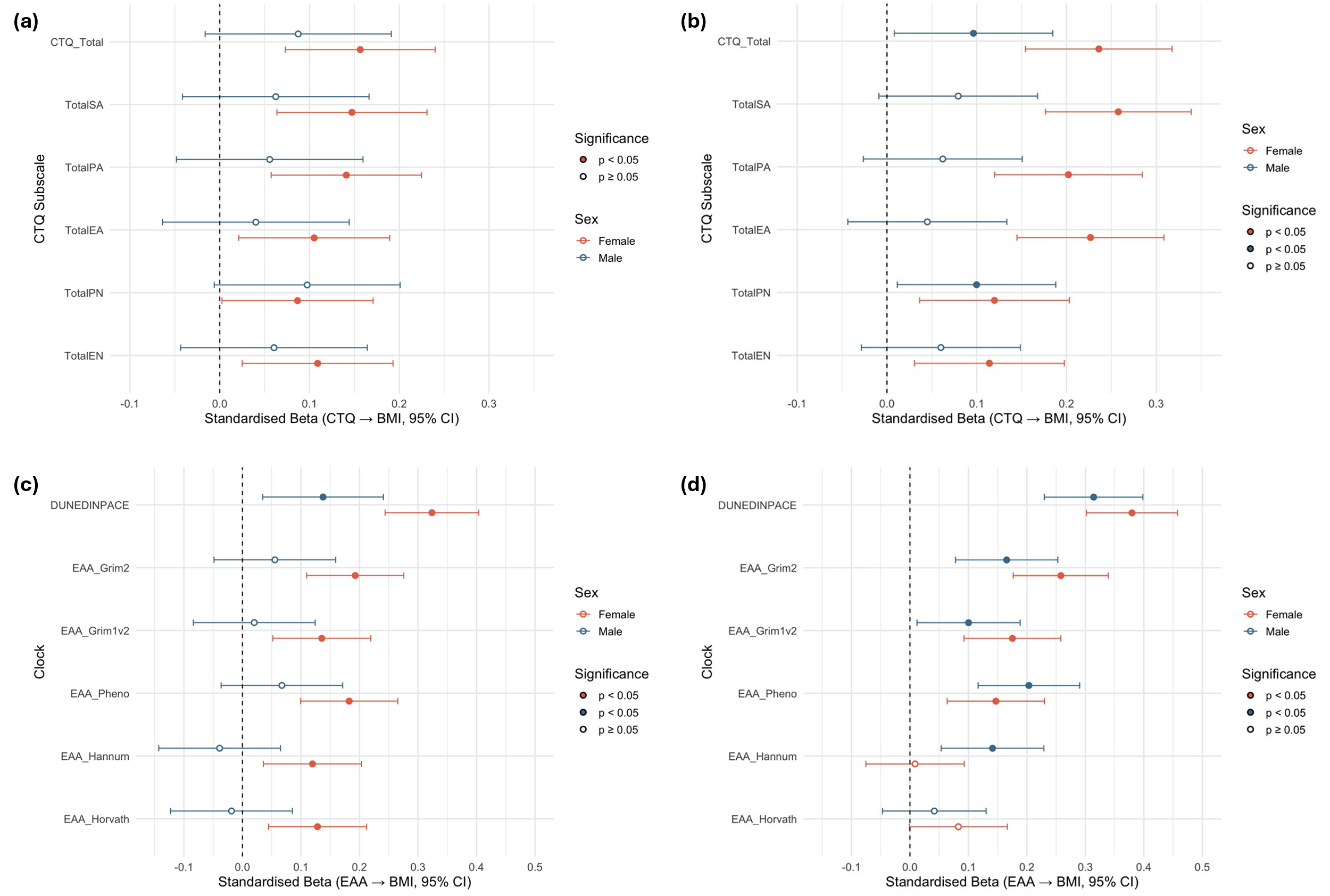

**Fig. S7.** Associations between CTQ and BMI and BMI and EAA. (a) CTQ and BMI in GS; (b) CTQ and BMI in MIDUS; (c) BMI and EAA in GS; (d) BMI and EAA in MIDUS. Stratified by sex. CTQ_Total is total CTQ score, TotalSA is total sexual abuse score, TotalPA is total physical abuse score, TotalEA is total emotional abuse score, TotalPN is total physical neglect score, and TotalEN is total emotional neglect score. EAA is epigenetic age acceleration, and CTQ is childhood trauma questionnaire. BMI was pre-calculated in both cohorts as weight divided by height squared. The results were used in analyses as a continuous score. In case of missing BMI data, mean imputation was employed.

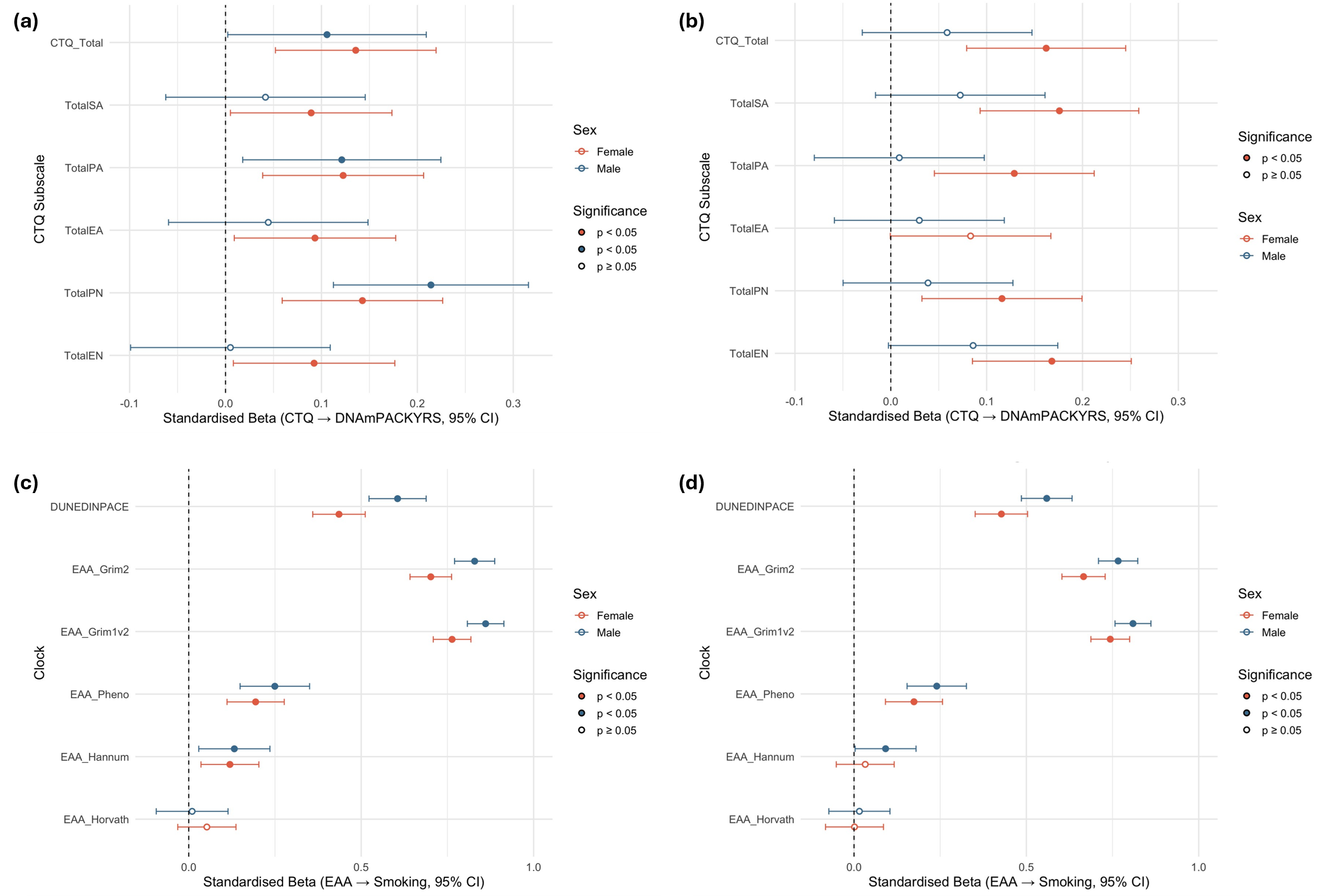

**Fig. S8.** Associations between CTQ and smoking and smoking and EAA. Associations between (a) CTQ and smoking in GS; (b) CTQ and smoking in MIDUS; (c) smoking and EAA in GS; (d) smoking and EAA in MIDUS. In all cases, the DNA methylation-based proxy, DNAmPACKYRS was used to determine smoking for participants. Stratified by sex. CTQ_Total is total CTQ score, TotalSA is total sexual abuse score, TotalPA is total physical abuse score, TotalEA is total emotional abuse score, TotalPN is total physical neglect score, and Total EN is total emotional neglect score. EAA is epigenetic age acceleration, and CTQ is childhood trauma questionnaire.

**Table S1**. Childhood trauma questionnaire categories

|  | **GS** | | | **MIDUS** | | |
| --- | --- | --- | --- | --- | --- | --- |
| **CTQ** | Female | Male | Total | Female | Male | Total |
| Total CTQ (mean (SD))  Range | 34.6 (13.3)  25-98 | 33.4 (9.2)  25-90 | 34.1 (11.8)  25-98 | 39.5 (15.8)  25-114 | 36.3 (11.2)  25-83 | 38.0 (13.9)  25-114 |
| Total SA (mean (SD))  Range | 6.4 (4.2)  5-25 | 5.4 (1.8)  5-21 | 6.0 (3.5)  5-25 | 7.0 (3.3)  5-25 | 6.9 (2.7)  5-20 | 6.9 (3.0)  5-25 |
| Total PA (mean (SD))  Range | 6.1 (2.6)  5-25 | 6.2 (2.3)  5-25 | 6.1 (2.5)  5-25 | 8.6 (4.5)  5-25 | 7.5 (3.4)  5-22 | 8.0 (4.1)  5-25 |
| Total EA (mean (SD))  Range | 7.5 (4.0)  5-25 | 6.4 (2.5)  5-25 | 7.1 (3.5)  5-25 | 7.4 (4.8)  5-25 | 5.6 (2.0)  5-23 | 6.5 (3.9)  5-25 |
| Total PN (mean (SD))  Range | 6.3 (2.4)  5-21 | 6.5 (2.3)  5-19 | 6.3 (2.4)  5-21 | 9.8 (4.6)  5-25 | 9.6 (4.2)  5-24 | 9.7 (4.4)  5-25 |
| Total EN (mean (SD))  Range | 8.4 (4.3)  5-25 | 8.8 (4.0)  5-24 | 8.5 (4.2)  5-25 | 6.8 (2.7)  5-20 | 6.7 (2.5)  5-18 | 6.7 (2.6)  5-20 |

Childhood trauma questionnaire (CTQ) scores with mean, standard deviation (SD) and ranges. Data are self-reported by participants of Generation Scotland (GS) and Midlife in the United States (MIDUS) cohorts. For individual categories the lowest possible score is 5 and maximum is 25. The minimum and maximum possible cumulative (total) scores are 25 and 125, respectively. Total SA is total sexual abuse score, Total PA is total physical abuse score, Total EA is total emotional abuse score, Total PN is total physical neglect score, and Total EN is total emotional neglect score.

**Table S2.** Associations between childhood trauma and chronic pain

|  | | **Model 1**  **OR (95% CI), p-value** | | | **Model 2**  **OR (95% CI), p-value** | | | **Model 3**  **OR (95% CI), p-value** | | |
| --- | --- | --- | --- | --- | --- | --- | --- | --- | --- | --- |
| **Variable** | **Sex** | **GS** | **MIDUS** | **Pooled** | **GS** | **MIDUS** | **Pooled** | **GS** | **MIDUS** | **Pooled** |
| CTQ_Total | F | 1.69 (1.39, 2.10), <0.001 | 1.49 (1.25, 1.80), <0.001 | 1.57 (1.37, 1.81), <0.001 | 1.66 (1.36, 2.07), <0.001 | 1.41 (1.17, 1.71), <0.001 | 1.52 (1.32, 1.75), <0.001 | 1.64 (1.34, 2.04), <0.001 | 1.34 (1.11, 1.63), 0.003 | 1.47 (1.27, 1.69), <0.001 |
|  | M | 1.28 (1.03, 1.61), 0.026 | 1.27 (1.05, 1.54), 0.013 | 1.27 (1.10, 1.47), 0.001 | 1.26 (1.01, 1.59), 0.037 | 1.27 (1.05, 1.53), 0.016 | 1.27 (1.10, 1.46), 0.001 | 1.25 (1.01, 1.57), 0.043 | 1.26 (1.04, 1.52), 0.020 | 1.26 (1.09, 1.45), 0.002 |
|  | C | 1.53 (1.32–1.79), <0.001 | 1.39 (1.22–1.59), <0.001 | 1.45 (1.31, 1.60), <0.001 | 1.49 (1.29–1.75), <0.001 | 1.35 (1.18–1.55), <0.001 | 1.41 (1.27, 1.56), <0.001 | 1.48 (1.27–1.73), <0.001 | 1.31 (1.15–1.51), <0.001 | 1.38 (1.25, 1.53), <0.001 |
| TotalEA | F | 1.62 (1.34, 1.99), <0.001 | 1.23 (1.03, 1.47), 0.020 | 1.36 (1.19, 1.56), <0.001 | 1.60 (1.32, 1.97), <0.001 | 1.16 (0.96, 1.39), 0.115 | 1.47 (1.30, 1.66), <0.001 | 1.59 (1.31, 1.95), <0.001 | 1.13 (0.93, 1.36), 0.210 | 1.25 (1.09, 1.43), 0.001 |
|  | M | 1.23 (0.99, 1.53), 0.065 | 1.21 (0.93, 1.35), 0.228 | 1.19 (1.03, 1.38), 0.022 | 1.22 (0.98, 1.53), 0.074 | 1.12 (0.93, 1.35), 0.237 | 1.16 (1.01, 1.34), 0.042 | 1.21 (0.97, 1.52), 0.088 | 1.11 (0.92, 1.35), 0.257 | 1.18 (1.01, 1.37), 0.032 |
|  | C | 1.48 (1.28–1.72), <0.001 | 1.19 (1.04–1.35), 0.009 | 1.31 (1.19, 1.44), <0.001 | 1.46 (1.26–1.70), <0.001 | 1.15 (1.01–1.31), 0.041 | 1.27 (1.16, 1.41), <0.001 | 1.44 (1.25–1.68), <0.001 | 1.13 (0.99–1.29), 0.076 | 1.26 (1.14, 1.39), <0.001 |
| TotalEN | F | 1.73 (1.43, 2.11), <0.001 | 1.32 (1.10, 1.58), 0.002 | 1.42 (1.24, 1.63), <0.001 | 1.71 (1.41, 2.09), <0.001 | 1.29 (1.08, 1.55), 0.006 | 1.47 (1.28, 1.68), <0.001 | 1.69 (1.40, 2.07), <0.001 | 1.22 (1.02, 1.47), 0.033 | 1.32 (1.15, 1.52), <0.001 |
|  | M | 1.25 (1.00, 1.56), 0.048 | 1.27 (1.05, 1.54), 0.012 | 1.19 (1.03, 1.38), 0.018 | 1.24 (0.99, 1.55), 0.057 | 1.27 (1.05, 1.53), 0.014 | 1.26 (1.09, 1.45), 0.002 | 1.24 (0.99, 1.55), 0.054 | 1.26 (1.04, 1.52), 0.017 | 1.18 (1.02, 1.36), 0.030 |
|  | C | 1.50 (1.30–1.73), <0.001 | 1.30 (1.14–1.48), <0.001 | 1.39 (1.26, 1.53), <0.001 | 1.48 (1.29–1.71), <0.001 | 1.28 (1.12–1.46), <0.001 | 1.37 (1.24, 1.51), <0.001 | 1.48 (1.28–1.71), <0.001 | 1.25 (1.10–1.42), <0.001 | 1.35 (1.22, 1.48), <0.001 |
| TotalPA | F | 1.31 (1.08, 1.63), 0.005 | 1.52 (1.27, 1.83), <0.001 | 1.39 (1.22, 1.59), <0.001 | 1.28 (1.05, 1.59), 0.011 | 1.45 (1.21, 1.76), <0.001 | 1.37 (1.19, 1.58), <0.001 | 1.26 (1.04, 1.57), 0.017 | 1.38 (1.14, 1.68), <0.001 | 1.39 (1.19, 1.62), <0.001 |
|  | M | 1.23 (0.99, 1.57), 0.060 | 1.17 (0.97, 1.42), 0.105 | 1.22 (1.06, 1.40), 0.006 | 1.21 (0.98, 1.53), 0.080 | 1.17 (0.96, 1.41), 0.117 | 1.19 (1.03, 1.37), 0.021 | 1.20 (0.97, 1.52), 0.096 | 1.16 (0.95, 1.40), 0.135 | 1.16 (1.01, 1.35), 0.042 |
|  | C | 1.28 (1.10–1.49), <0.001 | 1.36 (1.19–1.55), <0.001 | 1.32 (1.20, 1.46), <0.001 | 1.25 (1.08–1.46), 0.002 | 1.32 (1.16–1.51), <0.001 | 1.29 (1.17, 1.42), <0.001 | 1.23 (1.07–1.44), 0.004 | 1.29 (1.13–1.47), <0.001 | 1.26 (1.14, 1.39), <0.001 |
| TotalPN | F | 1.35 (1.13, 1.63), <0.001 | 1.33 (1.11, 1.59), 0.002 | 1.34 (1.18, 1.52), <0.001 | 1.34 (1.11, 1.62), 0.002 | 1.29 (1.08, 1.55), 0.006 | 1.31 (1.15, 1.50), <0.001 | 1.31 (1.10, 1.59), 0.003 | 1.25 (1.04, 1.51), 0.019 | 1.28 (1.12, 1.46), <0.001 |
|  | M | 1.04 (0.83, 1.30), 0.729 | 1.24 (1.02, 1.50), 0.028 | 1.15 (0.99, 1.33), 0.060 | 1.02 (0.82, 1.28), 0.832 | 1.23 (1.01, 1.49), 0.035 | 1.13 (0.98, 1.31), 0.092 | 1.01 (0.80, 1.27), 0.937 | 1.22 (1.01, 1.48), 0.040 | 1.13 (0.98, 1.31), 0.104 |
|  | C | 1.21 (1.06–1.39), 0.005 | 1.28 (1.13–1.46), <0.001 | 1.25 (1.14, 1.37), <0.001 | 1.20 (1.04–1.37), 0.011 | 1.26 (1.11–1.44), <0.001 | 1.23 (1.12, 1.35), <0.001 | 1.18 (1.03–1.36), 0.020 | 1.24 (1.09–1.41), 0.002 | 1.21 (1.10, 1.33), <0.001 |
| TotalSA | F | 1.26 (1.05, 1.52), 0.012 | 1.48 (1.23, 1.80), <0.001 | 1.50 (1.31, 1.71), <0.001 | 1.23 (1.03, 1.49), 0.022 | 1.39 (1.15, 1.70), <0.001 | 1.30 (1.14, 1.49), <0.001 | 1.21 (1.01, 1.47), 0.034 | 1.30 (1.07, 1.59), 0.009 | 1.46 (1.29, 1.65), <0.001 |
|  | M | 1.20 (0.97, 1.54), 0.099 | 1.18 (0.98, 1.43), 0.085 | 1.25 (1.08, 1.45), 0.002 | 1.19 (0.95, 1.54), 0.123 | 1.17 (0.97, 1.42), 0.097 | 1.18 (1.01, 1.37), 0.032 | 1.19 (0.95, 1.53), 0.133 | 1.17 (0.96, 1.41), 0.117 | 1.25 (1.08, 1.45), 0.002 |
|  | C | 1.23 (1.07–1.43), 0.003 | 1.33 (1.17–1.52), <0.001 | 1.28 (1.17, 1.42), <0.001 | 1.21 (1.05–1.40), 0.009 | 1.29 (1.13–1.48), <0.001 | 1.25 (1.13, 1.38), <0.001 | 1.19 (1.04–1.39), 0.014 | 1.25 (1.09–1.43), 0.001 | 1.22 (1.11, 1.35), <0.001 |

Relationships between childhood trauma questionnaire (CTQ) scores and chronic pain. The data are odds ratios (OR) with confidence intervals (CI) for reporting chronic pain per SD increase in CTQ score. CTQ Total is the cumulative score. TotalEA is the total score for emotional abuse; EN is emotional neglect; PA is physical abuse; PN is physical neglect; SA is sexual abuse. F is female, M is male and C is combined (male and female).

**Table S3.** Severity-dependent associations between childhood trauma and chronic pain

| **Variable** | **Sex** | **Model 1**  **OR (95% CI), p-value** | **Model 2**  **OR (95% CI), p-value** | **Model 3**  **OR (95% CI), p-value** |
| --- | --- | --- | --- | --- |
| Low | F | 1.80 (1.24, 2.61), <0.001 | 1.74 (1.20, 2.53), 0.001 | 1.73 (1.19, 2.53), 0.002 |
|  | M | 1.22 (0.91, 1.83), 0.518 | 1.21 (0.81, 1.82), 0.529 | 1.20 (0.79, 1.80), 0.592 |
|  | C | 1.496 (1.141, 1.961), 0.001 | 1.465 (1.116, 1.924), 0.003 | 1.438 (1.093, 1.892), 0.005 |
| Moderate | F | 1.93 (1.11, 3.38), 0.015 | 1.77 (1.01, 3.12), 0.047 | 1.72 (0.98, 3.05), 0.065 |
|  | M | 1.98 (0.95, 4.15), 0.078 | 1.83 (0.87, 3.85), 0.144 | 1.85 (0.87, 3.89), 0.136 |
|  | C | 1.954 (1.254, 3.045), 0.001 | 1.798 (1.148, 2.816), 0.006 | 1.781 (1.134, 2.797), 0.007 |
| Severe | F | 3.85 (1.99, 7.45), <0.001 | 3.51 (1.80, 6.83), <0.001 | 3.07 (1.57, 6.05), <0.001 |
|  | M | 3.76 (1.10, 12.86), 0.031 | 3.69 (1.08, 12.62), 0.034 | 3.48 (1.01, 12.00), 0.047 |
|  | C | 3.756 (2.110, 6.689), <0.001 | 3.484 (1.947, 6.234), <0.001 | 3.149 (1.751, 5.663), <0.001 |

Relationships between cumulative childhood trauma severity and chronic pain using combined data from GS and MIDUS. The data are odds ratios (OR) with confidence intervals (CI) for reporting chronic pain per SD increase in CTQ score for each category (see Methods). CTQ Total is the cumulative score. TotalEA is the total score for emotional abuse; EN is emotional neglect; PA is physical abuse; PN is physical neglect; SA is sexual abuse. F is female, M is male and C is combined (male and female).

**Table S4.** Associations between childhood trauma and EAA

|  | | **Model 1 - Std β (95% CI), p-value** | | | **Model 2 - Std β (95% CI), p-value** | | | **Model 3 - Std β (95% CI), p-value** | | |
| --- | --- | --- | --- | --- | --- | --- | --- | --- | --- | --- |
| **Variable** | **Sex** | **GS** | **MIDUS** | **Pooled** | **GS** | **MIDUS** | **Pooled** | **GS** | **MIDUS** | **Pooled** |
| GrimAge1v2 | F | 0.153 (0.073, 0.233), <0.001 | 0.168 (0.090, 0.247), <0.001 | 0.161 (0.105, 0.217), <0.001 | 0.140 (0.060, 0.221), <0.001 | 0.153 (0.073, 0.233), <0.001 | 0.147 (0.090, 0.203), <0.001 | 0.039 (-0.011, 0.088), 0.127 | 0.064 (0.010, 0.118), 0.020 | 0.050 (0.014, 0.087), 0.007 |
|  | M | 0.086 (-0.012, 0.184), 0.086 | 0.061 (-0.021, 0.143), 0.146 | 0.071 (0.008, 0.134), 0.026 | 0.088 (-0.010, 0.187), 0.078 | 0.057 (-0.026, 0.139), 0.179 | 0.070 (0.007, 0.133), 0.031 | 0.010 (-0.041, 0.060), 0.699 | 0.017 (-0.033, 0.068), 0.502 | 0.014 (-0.022, 0.049), 0.459 |
|  | C | 0.13 (0.07,0.18), 0.001 | 0.11 (0.06, 0.17), <0.001 | 0.13 (0.09, 0.16), <0.001 | 0.12 (0.06, 0.18), <0.001 | 0.11 (0.05, 0.16), <0.001 | 0.11 (0.07, 0.16), <0.001 | 0.03 (-0.01, 0.06), 0.135 | 0.04 (0.00, 0.08), 0.027 | 0.03 (0.00, 0.06), 0.011 |
| GrimAge2 | F | 0.150 (0.072, 0.229), <0.001 | 0.172 (0.095, 0.249), <0.001 | 0.161 (0.106, 0.216), <0.001 | 0.130 (0.052, 0.208), 0.001 | 0.139 (0.062, 0.216), <0.001 | 0.135 (0.080, 0.189), <0.001 | 0.037 (-0.015, 0.090), 0.165 | 0.061 (0.004, 0.118), 0.036 | 0.048 (0.009, 0.087), 0.015 |
|  | M | 0.095 (-0.002, 0.192), 0.054 | 0.074 (-0.007, 0.155), 0.072 | 0.083 (0.020, 0.145), 0.009 | 0.095 (-0.003, 0.192), 0.056 | 0.064 (-0.017, 0.144), 0.120 | 0.077 (0.014, 0.139), 0.016 | 0.019 (-0.034, 0.072), 0.486 | 0.028 (-0.026, 0.081), 0.311 | 0.023 (-0.014, 0.061), 0.222 |
|  | C | 0.13 (0.07,0.19), <0.001 | 0.10 (0.07,0.18), <0.001 | 0.13 (0.09,0.17), <0.001 | 0.12 (0.06,0.18), <0.001 | 0.10 (0.05, 0.16), <0.001 | 0.11 (0.07, 0.15), <0.001 | 0.03 (-0.01, 0.07), 0.146 | 0.04 (0.01,0.08), 0.027 | 0.04 (0.00, 0.06), 0.008 |
| DunedinPACE | F | 0.116 (0.036, 0.195), 0.004 | 0.141 (0.064, 0.219), <0.001 | 0.129 (0.073, 0.184), <0.001 | 0.073 (-0.003, 0.149), 0.059 | 0.079 (0.005, 0.153), 0.036 | 0.076 (0.023, 0.129), 0.005 | 0.021 (-0.048, 0.090), 0.549 | 0.023 (-0.047, 0.092), 0.521 | 0.022 (-0.027, 0.071), 0.379 |
|  | M | 0.112 (0.018, 0.206), 0.020 | 0.111 (0.033, 0.189), 0.005 | 0.111 (0.051, 0.171), <0.001 | 0.104 (0.011, 0.197), 0.019 | 0.086 (0.011, 0.161), 0.025 | 0.093 (0.035, 0.151), 0.002 | 0.057 (-0.021, 0.136), 0.152 | 0.050 (-0.018, 0.118), 0.149 | 0.053 (0.002, 0.104), 0.043 |
|  | C | 0.11 (0.05, 0.17), <0.001 | 0.14 (0.09, 0.20), <0.001 | 0.11 (0.07, 0.16), <0.001 | 0.08 (0.02, 0.14), 0.005 | 0.09 (0.03, 0.14), 0.012 | 0.09 (0.04, 0.13), <0.001 | 0.03 (-0.02, 0.08), 0.231 | 0.04 (-0.01, 0.09), 0.132 | 0.03 (-0.01, 0.07), 0.096 |
| PhenoAge | F | 0.066 (-0.011, 0.143), 0.092 | 0.095 (0.016, 0.174), 0.019 | 0.080 (0.025, 0.135), 0.004 | 0.047 (-0.030, 0.124), 0.231 | 0.064 (-0.016, 0.143), 0.115 | 0.055 (0.000, 0.111), 0.050 | 0.023 (-0.053, 0.099), 0.549 | 0.045 (-0.034, 0.124), 0.268 | 0.034 (-0.021, 0.088), 0.230 |
|  | M | 0.101 (0.002, 0.200), 0.045 | 0.070 (-0.011, 0.150), 0.088 | 0.082 (0.020, 0.145), 0.010 | 0.094 (-0.04, 0.193), 0.060 | 0.053 (-0.026, 0.133), 0.186 | 0.066 (0.000, 0.132), 0.049 | 0.070 (-0.025, 0.166), 0.149 | 0.046 (-0.033, 0.124), 0.253 | 0.056 (-0.005, 0.116), 0.072 |
|  | C | 0.08 (0.02,0.14), 0.012 | 0.08 (0.03, 0.14), 0.003 | 0.08 (0.04, 0.12), <0.001 | 0.06 (0.00, 0.12), 0.047 | 0.06 (0.00, 0.11), 0.043 | 0.06 (0.02, 0.10), 0.004 | 0.04 (-0.02, 0.10), 0.232 | 0.04 (-0.01, 0.10), 0.116 | 0.04 (0.00,0.08), 0.053 |
| Hannum | F | 0.045 (-0.035, 0.124), 0.270 | 0.061 (-0.016, 0.139), 0.119 | 0.053 (-0.002, 0.109), 0.060 |  | | | | | |
|  | M | 0.019 (-0.082, 0.119), 0.712 | 0.038 (-0.047, 0.123), 0.375 | 0.030 (-0.035, 0.095), 0.364 |  |  |  |  |  |  |
|  | C | 0.04 (-0.02, 0.10), 0.233 | 0.05 (-0.01, 0.11), 0.084 | 0.05 (0.00,0.09), 0.038 |  |  |  |  |  |  |
| Horvath | F | -0.019 (-0.103, 0.065), 0.655 | 0.021 (-0.064, 0.107), 0.623 | 0.001 (-0.059, 0.061), 0.983 |  |  |  |  |  |  |
|  | M | 0.002 (-0.099, 0.103), 0.966 | 0.058 (-0.030, 0.143), 0.188 | 0.034 (-0.031, 0.100), 0.306 |  |  |  |  |  |  |
|  | C | -0.01 (-0.08, 0.05), 0.717 | 0.04 (-0.02, 0.10), 0.212 | 0.02 (-0.03, 0.06), 0.450 |  |  |  |  |  |  |

Relationships between childhood trauma and EAA established using the DNA methylation age clocks. F is female, M is male and C is combined (male and female). Sequential Models 2 and 3 are not presented for Hannum and Horvath because of no consistent associations in Model 1.

**Table S5.** Severity-dependent associations between childhood trauma and EAA

| **Variable** | **Sex** | **ACE score (vs none)** | **Model 1**  **Mean EAA (95% CI), p-value** | **Model 2**  **Mean EAA (95% CI), p-value** | **Model 3**  **Mean EAA (95% CI), p-value** |
| --- | --- | --- | --- | --- | --- |
| GrimAge1v2 | F | Low | 0.674 (0.009, 1.339), 0.046 | 0.614 (-0.051, 1.278), 0.078 | 0.466 (0.038, 0.895), 0.029 |
|  |  | Moderate | 1.215 (0.209, 2.222), 0.013 | 1.065 (0.056, 2.075), 0.036 | 0.681 (0.029, 1.333), 0.038 |
|  |  | Severe | 2.410 (1.294, 3.525), <0.001 | 2.229 (1.109, 3.350), <0.001 | 0.870 (0.142, 1.598), 0.014 |
|  | M | Low | 0.456 (-0.323, 1.235), 0.373 | 0.455 (-0.324, 1.234), 0.376 | 0.026 (-0.465, 0.517), 0.994 |
|  |  | Moderate | -0.251 (-1.723, 1.220), 0.932 | -0.280 (-1.762, 1.203), 0.917 | -0.064 (-0.997, 0.868), 0.990 |
|  |  | Severe | 1.713 (-0.658, 4.083), 0.216 | 1.705 (-0.667, 4.077), 0.219 | 0.039 (-1.457, 1.535), 0.999 |
|  | C | Low | 0.626 (0.116, 1.135), 0.011 | 0.594 (0.085, 1.104), 0.017 | 0.291 (-0.045, 0.627), 0.107 |
|  |  | Moderate | 0.611 (-0.234, 1.455), 0.216 | 0.497 (-0.352, 1.346), 0.374 | 0.426 (-0.133, 0.985), 0.180 |
|  |  | Severe | 2.162 (1.124, 3.200), <0.001 | 2.048 (1.007, 3.088), <0.001 | 0.895 (0.208, 1.583), 0.006 |
| GrimAge2 | F | Low | 0.620 (-0.109, 1.348), 0.116 | 0.491 (-0.229, 1.210), 0.257 | 0.343 (-0.166, 0.852), 0.265 |
|  |  | Moderate | 1.364 (0.262, 2.466), 0.010 | 1.043 (-0.049, 2.136), 0.065 | 0.658 (-0.115, 1.432), 0.116 |
|  |  | Severe | 2.653 (1.431, 3.875), <0.001 | 2.269 (1.055, 3.482), <0.001 | 0.906 (0.043, 1.770), 0.037 |
|  | M | Low | 0.608 (-0.242, 1.457), 0.223 | 0.599 (-0.249, 1.446), 0.232 | 0.136 (-0.404, 0.677), 0.852 |
|  |  | Moderate | 0.403 (-1.202, 2.008), 0.853 | 0.223 (-1.390, 1.836), 0.956 | 0.455 (-0.571, 1.482), 0.581 |
|  |  | Severe | 2.284 (-0.302, 4.871), 0.098 | 2.236 (-0.345, 4.817), 0.107 | 0.438 (-1.210, 2.085), 0.835 |
|  | C | Low | 0.662 (0.108, 1.216), 0.014 | 0.588 (0.038, 1.138), 0.032 | 0.274 (-0.107, 0.655), 0.219 |
|  |  | Moderate | 0.976 (0.058, 1.895), 0.034 | 0.710 (-0.205, 1.626), 0.169 | 0.637 (0.002, 1.271), 0.049 |
|  |  | Severe | 2.514 (1.685, 3.642), <0.001 | 2.247 (1.124, 3.370), <0.001 | 1.055 (0.275, 1.835), 0.004 |
| DunedinPACE | F | Low | 0.020 (0.001, 0.040), 0.032 | 0.014 (-0.004, 0.032), 0.162 | 0.012 (-0.005, 0.029), 0.208 |
|  |  | Moderate | 0.037 (0.008, 0.066), 0.008 | 0.022 (-0.006, 0.049), 0.161 | 0.014 (-0.011, 0.040), 0.420 |
|  |  | Severe | 0.068 (0.036, 0.100), <0.001 | 0.049 (0.019, 0.080), <0.001 | 0.030 (0.001, 0.058), 0.039 |
|  | M | Low | 0.026 (0.006, 0.047), 0.008 | 0.026 (0.005, 0.046), 0.008 | 0.017 (-0.001, 0.035), 0.073 |
|  |  | Moderate | 0.057 (0.018, 0.096), 0.002 | 0.044 (0.005, 0.082), 0.022 | 0.044 (0.011, 0.078), 0.006 |
|  |  | Severe | 0.072 (0.009, 0.136), 0.021 | 0.068 (0.006, 0.130), 0.026 | 0.032 (-0.022, 0.087), 0.364 |
|  | C | Low | 0.024 (0.010, 0.038), <0.001 | 0.020 (0.006, 0.052), 0.001 | 0.015 (0.002, 0.027), 0.013 |
|  |  | Moderate | 0.044 (0.020, 0.067), <0.001 | 0.029 (0.006, 0.052), 0.007 | 0.025 (0.005, 0.046), 0.011 |
|  |  | Severe | 0.064 (0.035, 0.093), <0.001 | 0.049 (0.021, 0.077), <0.001 | 0.028 (0.003, 0.054), 0.023 |
| PhenoAge | F | Low | 0.933 (-0.063, 1.929), 0.072 | 0.799 (-0.191, 1.789), 0.145 | 0.760 (-0.222, 1.741), 0.170 |
|  |  | Moderate | 1.250 (-0.257, 2.756), 0.130 | 0.918 (-0.587, 2.422), 0.342 | 0.815 (-0.677, 2.307), 0.428 |
|  |  | Severe | 3.195 (1.525, 4.866), <0.001 | 2.797 (1.127, 4.466), <0.001 | 2.434 (0.769, 4.100), 0.002 |
|  | M | Low | 1.222 (0.187, 2.256), 0.015 | 1.200 (0.177, 2.222), 0.016 | 1.058 (0.053, 2.063), 0.036 |
|  |  | Moderate | 2.226 (0.272, 4.181), 0.020 | 1.782 (-0.164, 3.727), 0.082 | 1.853 (-0.056, 3.762), 0.060 |
|  |  | Severe | 3.143 (-0.006, 6.292), 0.051 | 3.023 (-0.091, 6.136), 0.060 | 2.473 (-0.591, 5.536), 0.145 |
|  | C | Low | 1.112 (0.398, 1.827), <0.001 | 1.012 (0.305, 1.721), 0.002 | 0.923 (0.223, 1.622), 0.005 |
|  |  | Moderate | 1.591 (0.407, 2.775), 0.004 | 1.233 (0.054, 2.413), 0.038 | 1.212 (0.048, 2.377), 0.039 |
|  |  | Severe | 3.144 (1.690, 4.599), <0.001 | 2.786 (1.339, 4.232), <0.001 | 2.444 (1.012, 3.876), <0.001 |
| Hannum | F | Low | -0.094 (-0.734, 0.546), 0.950 |  | |
|  |  | Moderate | 0.249 (-0.719, 1.218), 0.845 |  |  |
|  |  | Severe | 0.614 (-0.460, 1.687), 0.393 |  |  |
|  | M | Low | 0.672 (-0.098, 1.442), 0.104 |  |  |
|  |  | Moderate | -0.182 (-1.638, 1.273), 0.964 |  |  |
|  |  | Severe | -0.050 (-2.395, 2.296), 0.999 |  |  |
|  | C | Low | 0.281 (-0.213, 0.775), 0.397 |  |  |
|  |  | Moderate | 0.074 (-0.745, 0.893), 0.982 |  |  |
|  |  | Severe | 0.352 (-0.654, 1.359), 0.722 |  |  |
| Horvath | F | Low | -0.308 (-1.005, 0.390), 0.585 |  |  |
|  |  | Moderate | 0.059 (-0.996, 1.113), 0.993 |  |  |
|  |  | Severe | 0.453 (-0.716, 1.622), 0.666 |  |  |
|  | M | Low | 0.922 (0.154, 1.691), 0.013 |  |  |
|  |  | Moderate | 0.106 (-1.347, 1.558), 0.988 |  |  |
|  |  | Severe | 0.688 (-1.652, 3.028), 0.799 |  |  |
|  | C | Low | 0.334 (-0.185, 0.853), 0.300 |  |  |
|  |  | Moderate | 0.065 (-0.795, 0.925), 0.987 |  |  |
|  |  | Severe | 0.371 (-0.686, 1.428), 0.721 |  |  |

Relationships between childhood trauma severity categories (see Methods) and EAA established using the DNA methylation age clocks. F is female, M is male and C is combined (male and female). Sequential Models 2 and 3 are not presented for Hannum and Horvath because of no consistent associations in Model 1.

**Table S6.** Associations between EAA and chronic pain

|  | | **Model 1 - OR (95% CI), p-value** | | | **Model 2 - OR (95% CI), p-value** | | | **Model 3 - OR (95% CI), p-value** | | |
| --- | --- | --- | --- | --- | --- | --- | --- | --- | --- | --- |
| **Variable** | **Sex** | **GS** | **MIDUS** | **Pooled** | **GS** | **MIDUS** | **Pooled** | **GS** | **MIDUS** | **Pooled** |
| GrimAge1v2 | F | 1.23 (1.03, 1.48), 0.025 | 1.37 (1.13, 1.66), 0.001 | 1.29 (1.13, 1.48), <0.001 | 1.21 (1.01, 1.56), 0.042 | 1.33 (1.10, 1.61), 0.004 | 1.28 (1.11, 1.47), <0.001 | 1.04 (0.76, 1.40), 0.822 | 0.73 (0.53, 0.99), 0.043 | 0.87 (0.70, 1.09), 0.230 |
|  | M | 1.04 (0.82, 1.31), 0.759 | 1.05 (0.85, 1.29), 0.633 | 1.05 (0.89, 1.22), 0.575 | 1.05 (0.83, 1.33), 0.690 | 1.04 (0.85, 1.28), 0.685 | 1.04 (0.89, 1.22), 0.583 | 0.69 (0.42, 1.11), 0.130 | 0.78 (0.55, 1.12), 0.182 | 0.75 (0.56, 1.00), 0.047 |
|  | C | 1.16 (1.00, 1.34), 0.049 | 1.20 (1.04, 1.38), 0.010 | 1.19 (1.07, 1.31), 0.001 | 1.14 (0.99, 1.32), 0.077 | 1.18 (1.02, 1.35), 0.024 | 1.16 (1.05, 1.28), 0.004 | 0.89 (0.68, 1.16), 0.386 | 0.74 (0.58, 0.93), 0.012 | 0.80 (0.67, 0.96), 0.015 |
| GrimAge2 | F | 1.19 (0.99, 1.44), 0.063 | 1.34 (1.10, 1.63), 0.003 | 1.26 (1.10, 1.44), 0.001 | 1.16 (0.96, 1.40), 0.119 | 1.25 (1.02, 1.53), 0.028 | 1.20 (1.05, 1.38), 0.009 | 0.95 (0.71, 1.26), 0.712 | 0.71 (0.53, 0.94), 0.018 | 0.82 (0.67, 1.01), 0.057 |
|  | M | 1.09 (0.86, 1.38), 0.464 | 1.05 (0.85, 1.29), 0.676 | 1.07 (0.91, 1.25), 0.414 | 1.09 (0.85, 1.38), 0.488 | 1.03 (0.83, 1.27), 0.789 | 1.06 (0.90, 1.24), 0.506 | 0.83 (0.53, 1.30), 0.417 | 0.80 (0.57, 1.12), 0.201 | 0.81 (0.62, 1.06), 0.127 |
|  | C | 1.15 (1.00, 1.34), 0.054 | 1.18 (1.03, 1.36), 0.019 | 1.17 (1.05, 1.29), 0.003 | 1.13 (0.97, 1.31), 0.116 | 1.13 (0.98, 1.31), 0.089 | 1.13 (1.02, 1.25), 0.022 | 0.89 (0.70, 1.13), 0.339 | 0.74 (0.60, 0.92), 0.007 | 0.80 (0.68, 0.94), 0.007 |
| DunedinPACE | F | 1.13 (0.94, 1.35), 0.203 | 1.13 (0.93, 1.37), 0.225 | 1.13 (0.99, 1.29), 0.070 |  | | | | | |
|  | M | 1.13 (0.88, 1.44), 0.330 | 1.09 (0.87, 1.36), 0.437 | 1.11 (0.94, 1.31), 0.225 |  |  |  |  |  |  |
|  | C | 1.13 (0.97, 1.31), 0.109 | 1.10 (0.95, 1.28), 0.184 | 1.11 (1.00, 1.24), 0.044 |  |  |  |  |  |  |
| PhenoAge | F | 0.73 (0.60, 0.88), 0.001 | 1.04 (0.86, 1.27), 0.658 | 0.87 (0.75, 1.00), 0.043 |  |  |  |  |  |  |
|  | M | 1.13 (0.89, 1.42), 0.318 | 1.16 (0.94, 1.44), 0.175 | 1.15 (0.98, 1.34), 0.089 |  |  |  |  |  |  |
|  | C | 0.87 (0.75, 1.01), 0.066 | 1.09 (0.95, 1.26), 0.222 | 0.98 (0.88, 1.09), 0.693 |  |  |  |  |  |  |
| Hannum | F | 0.85 (0.71, 1.03), 0.098 | 1.07 (0.87, 1.31), 0.510 | 0.94 (0.82, 1.08), 0.406 |  |  |  |  |  |  |
|  | M | 0.89 (0.70, 1.12), 0.326 | 1.00 (0.81, 1.23), 0.980 | 0.95 (0.81, 1.11), 0.518 |  |  |  |  |  |  |
|  | C | 0.86 (0.74, 1.00), 0.052 | 1.03 (0.89, 1.19), 0.645 | 0.94 (0.85, 1.05), 0.282 |  |  |  |  |  |  |
| Horvath | F | 0.89 (0.75, 1.06), 0.208 | 0.91 (0.76, 1.09), 0.292 | 0.90 (0.79, 1.02), 0.096 |  |  |  |  |  |  |
|  | M | 0.97 (0.77, 1.21), 0.764 | 1.13 (0.92, 1.38), 0.240 | 1.06 (0.91, 1.23), 0.482 |  |  |  |  |  |  |
|  | C | 0.92 (0.80, 1.06), 0.241 | 1.01 (0.88, 1.15), 0.920 | 0.97 (0.88, 1.06), 0.487 |  |  |  |  |  |  |

Relationships between EAA and chronic pain. F is female, M is male and C is combined (male and female). Sequential Models 2 and 3 are not presented for DunedinPACE, PhenoAge, Hannum and Horvath because there were no consistent associations between EAA and chronic pain in Model 1.

**Table S7.** Associations between childhood trauma and pain - mediation by EAA

|  | | **Model 1 - ACME (95% CI), p-value** | | | **Model 2 - ACME (95% CI), p-value** | | | **Model 3 - ACME (95% CI), p-value** | | |
| --- | --- | --- | --- | --- | --- | --- | --- | --- | --- | --- |
| **Variable** | **Sex** | **GS** | **MIDUS** | **Pooled** | **GS** | **MIDUS** | **Pooled** | **GS** | **MIDUS** | **Pooled** |
| GrimAge1v2 | F | 4.564e-03  (-0.001, 0.013), 0.142 | 1.011e-02 (0.003, 0.020), 0.004 | 6.805e-03 (0.001, 0.012), 0.014 | 3.838e-03  (-0.002, 0.011), 0.168 | 8.410e-03 (0.002, 0.017), 0.002 | 5.799e-03 (0.001, 0.011), 0.021 | 1.608e-03  (-0.002, 0.007), 0.454 | -2.809e-03  (-0.009, 0.002), 0.226 | 7.996e-05  (-0.003, 0.003), 0.961 |
|  | M | -1.256e-03  (-0.011, 0.008), 0.982 | 3.197e-04  (-0.005, 0.005), 0.858 | -2.210e-05  (-0.004, 0.004), 0.992 | 1.944e-04  (-0.009, 0.009), 0.966 | 2.215e-04  (-0.005, 0.005), 0.928 | 2.151e-04  (-0.004, 0.005), 0.923 | -9.288e-04  (-0.012, 0.005), 0.796 | -8.751e-04  (-0.006, 0.003), 0.680 | -8.869e-04  (-0.005, 0.003), 0.662 |
|  | C | 8.453e-04  (-3.214e-03, 5.321e-03), 0.676 | 4.878e-03 (5.054e-04, 1.035e-02), 0.030 | 2.576e-03 (6.487e-04, 5.800e-03), 0.117 | 4.709e-04  (-0.003, 0.004), 0.838 | 4.162e-03  (0.000, 0.009), 0.050 | 1.862e-03  (-0.001, 0.005), 0.186 | -5.028e-04  (-0.003, 0.001), 0.646 | -1.727e-03  (-0.006, 0.001), 0.492 | -8.041e-04  (-0.003, 0.001), 0.364 |
| GrimAge2 | F | 3.542e-03  (-0.003, 0.011), 0.248 | 9.801e-03 (0.002, 0.019), 0.010 | 6.071e-03 (0.001, 0.011), 0.028 | 2.577e-03  (-0.003, 0.011), 0.354 | 6.355e-03 (0.000, 0.014), 0.032 | 4.466e-03  (-0.000, 0.009), 0.077 | 3.613e-04  (-0.003, 0.005), 0.788 | -3.195e-03  (-0.010, 0.001), 0.116 | -0.342e-03  (-0.005, 0.002), 0.416 |
|  | M | 1.048e-03  (-0.006, 0.011), 0.746 | 1.821e-04 (-0.005, 0.005), 0.966 | 4.047e-04 (-0.004, 0.005), 0.854 | 9.394e-04  (-0.008, 0.010), 0.876 | -4.494e-05  (-0.005, 0.005), 0.940 | 1.872e-04  (-0.004, 0.005), 0.933 | -7.656e-04  (-0.009, 0.004), 0.808 | -1.411e-03  (-0.008, 0.002), 0.528 | -1.171e-03  (-0.005, 0.003), 0.562 |
|  | C | 1.429e-03  (-3.215e-03, 6.021e-03), 0.540 | 4.506e-03 (1.847e-05, 9.783e-03), 0.050 | 2.882e-03 (4.730e-04, 6.237e-03), 0.092 | 7.371e-04  (-0.003, 0.005), 0.750 | 2.914e-03  (-0.001, 0.007), 0.132 | 1.826e-03  (-0.001, 0.005), 0.206 | -3.519e-04  (-0.003, 0.002), 0.728 | -2.838e-03  (-0.007, 0.001), 0.162 | -1.050e-03  (-0.003, 0.001), 0.332 |
| DunedinPACE | F | 1.500e-03  (-0.003, 0.008), 0.534 | 1.558e-03  (-0.004, 0.008), 0.530 | 1.526e-03  (-0.003, 0.006), 0.461 |  | | | | | |
|  | M | 3.157e-03  (-0.006, 0.015), 0.516 | 1.330e-03  (-0.005, 0.008), 0.656 | 1.836e-03  (-0.004, 0.007), 0.515 |  |  |  |  |  |  |
|  | C | 1.168e-03  (-3.079e-03, 5.431e-03), 0.574 | 1.536e-03  (-2.907e-03, 6.141e-03), 0.492 | 1.341e-03 (1.758e-03, 4.439e-03), 0.396 |  |  |  |  |  |  |
| PhenoAge | F | -5.111e-03  (-0.012, 0.000), 0.056 | 1.083e-03  (-0.003, 0.006), 0.572 | -1.147e-03  (-0.005, 0.002), 0.532 |  |  |  |  |  |  |
|  | M | 2.322e-03  (-0.005, 0.013), 0.554 | 2.448e-03  (-0.002, 0.010), 0.318 | 2.409e-03  (-0.003, 0.007), 0.344 |  |  |  |  |  |  |
|  | C | -3.426e-03  (-7.902e-03, -4.257e-04), 0.028 | 1.689e-03  (-1.032e-03, 5.387e-03), 0.246 | -4.815e-04  (-2.917e-03, 1.954e-03), 0.698 |  |  |  |  |  |  |
| Hannum | F | -1.694e-03  (-0.006, 0.002), 0.358 | 1.109e-03  (-0.001, 0.006), 0.376 | -1.065e-04  (-0.003, 0.003), 0.937 |  |  |  |  |  |  |

|  | M | -7.405e-04  (-0.007, 0.005), 0.842 | -6.385e-05  (-0.004, 0.003), 0.900 | -2.356e-04  (-0.003, 0.003), 0.879 |
| --- | --- | --- | --- | --- |
|  | C | -1.402e-03  (-5.863e-03, 1.618e-03), 0.328 | 2.408e-04  (-9.967e-04, 2.321e-03), 0.690 | -2.920e-05  (-1.546e-03, 1.487e-03), 0.970 |
| Horvath | F | 4.679e-04  (-0.002, 0.003), 0.712 | -3.579e-04  (-0.003, 0.002), 0.750 | 5.500e-05  (-0.002, 0.002), 0.951 |
|  | M | -2.307e-05  (-0.005, 0.005), 0.944 | 1.548e-03  (-0.002, 0.008), 0.518 | 7.625e-04  (-0.003, 0.004), 0.673 |
|  | C | 4.145e-04  (-1.404e-03, 2.696e-03), 0.726 | -2.652e-05  (-1.550e-03, 1.199e-03), 0.922 | 1.103e-04  (-1.031e-03, 1.252e-03), 0.850 |

Mediation analysis examining whether EAA mediated associations between childhood trauma and chronic pain. F is female, M is male and C is combined (male and female). Sequential Models 2 and 3 are not presented for DunedinPACE, PhenoAge, Hannum and Horvath because there was no mediation in Model 1.
